# Variability in proteoglycan biosynthetic genes reveals new facets of heparan sulfates diversity. A systematic review and analysis

**DOI:** 10.1101/2022.04.18.22273971

**Authors:** Mohand Ouidir Ouidja, Denis S.F. Biard, Sandrine Chantepie, Xavier Laffray, Gael Le Douaron, Minh-Bao Huynh, Nicolas Rebergue, Auriane Maïza, Karla Rubio, Oscar González-Velasco, Guillermo Barreto, Javier De Las Rivas, Dulce Papy-Garcia

## Abstract

Proteoglycans are complex macromolecules formed of glycosaminoglycan chains covalently linked to core proteins through a linker tetrasaccharide common to heparan sulfate proteoglycans (HSPG) and chondroitin sulfate proteoglycans (CSPG). Biosynthesis of a single proteoglycan requires the expression of dozens of genes, which together create the large structural and functional diversity reflected by the numerous diseases or syndromes associated to their genetic variability. Among proteoglycans, HSPG are the most structurally and functionally complex. To decrease this complexity, we retrieved and linked information on pathogenic variants, polymorphism, expression, and literature databases for 50 genes involved in the biosynthesis of HSPG core proteins, heparan sulfate (HS) chains, and their linker tetrasaccharide. This resulted in a new gene organization and biosynthetic pathway representation in which the phenotypic continuum of disorders as linkeropathies and other pathologies could be predictable. Moreover, ubiquitous *NDST1, GLCE, HS2ST1*, and *HS6ST1* appeared to generate ubiquitous heparan sulfate (HS) sequences essential for normal development and homeostasis, whereas the tissue restricted *NDST2-4, HS6ST2-3*, and *HS3ST1-6* appeared to generate specialized HS sequences mainly involved in responsiveness to stimuli. Supported by data on genetic polymorphism and clinical variants, we afford a new vision of HSPG involvement in homeostasis, disease, vulnerability to disease, and behavioral disorders.

## 1. Introduction

Proteoglycans are a family of macromolecules involved in virtually all aspects of cell biology and physiology^1^. These complex molecules consist in a core protein carrying one or several glycosaminoglycan (GAG) chains, either chondroitin sulfates (CS), heparan sulfates (HS), keratan sulfates (KS), or combinations of them. In HS proteoglycans (HSPG) and CS proteoglycans (CSPG), the glycanic chain is connected to their core proteins trough a common GAG-core protein linker tetrasaccharide (GAG-CP) (Fig. 1)^1-3^. Both the identity of the core protein and the structure of the glycanic chain confer to proteoglycans a great functional diversity, as reflected by the multiplicity of diseases or syndromes resulting from defects on genes coding for their biosynthetic machinery. Among proteoglycans, HS proteoglycans (HSPG) display the greater structural and functional diversity. This is principally due to the diversity and differential expression of the numerous genes required for their synthesis (Fig. 1). Interestingly, whereas various of these genes are essential for normal development and homeostasis, several others are not, suggesting that some, but not all, HSPG play essential biological roles depending on the core protein and associated HS chains generated in each tissue and cell by its HSPG biosynthetic machinery.

**Fig. 1.**
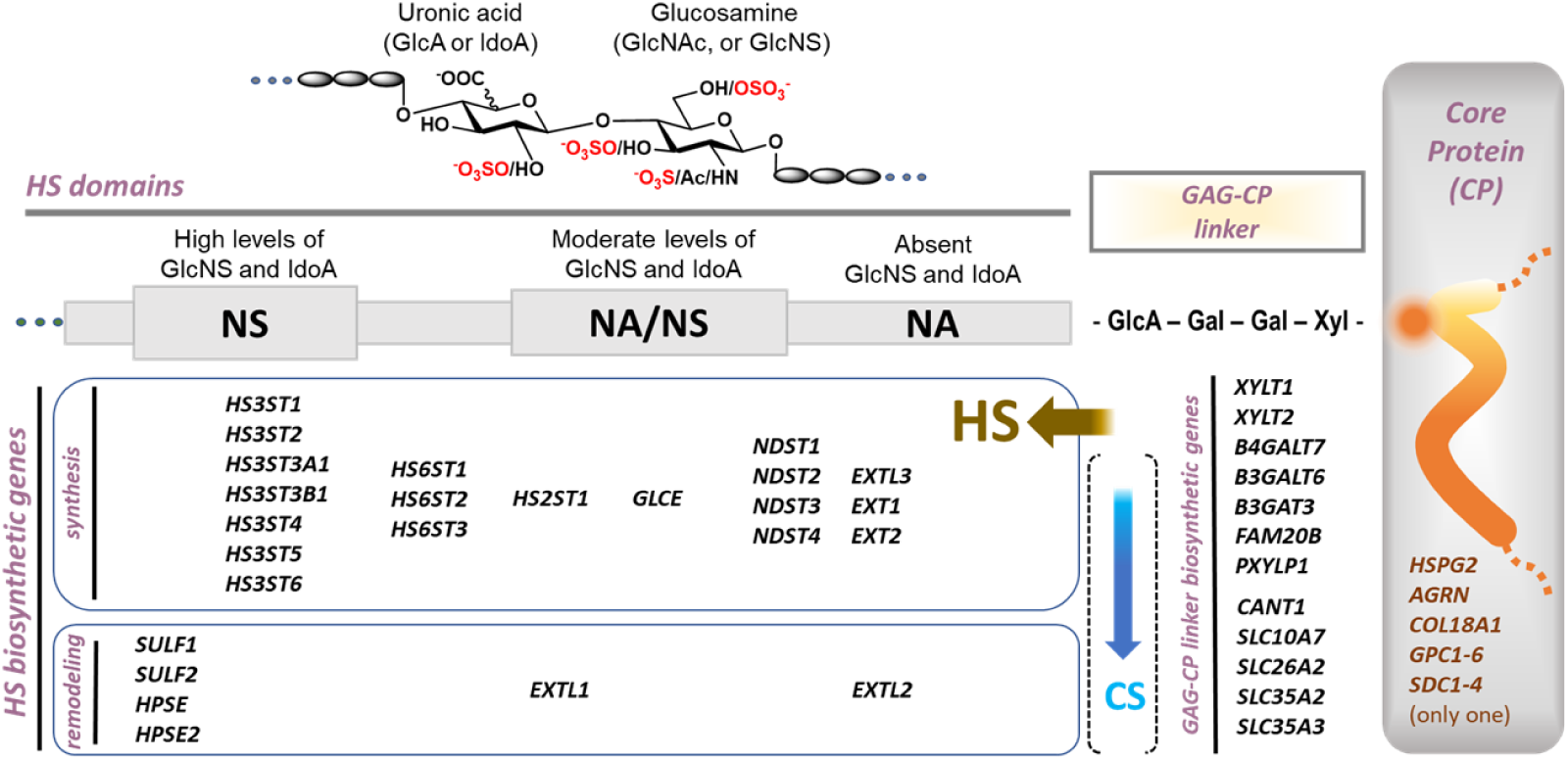
Schematic representation of a HSPG and their biosynthetic genes. HSPG are made of a core protein (CP) carrying a glycosaminoglycan-CP linker tetrasaccharide (GAG-CP linker) in which either heparan sulfate (HS) or chondroitin sulfate (CS) chains are elongated. The HS chain is formed by constitutive disaccharides composed of a uronic acid (GlcA or IdoA) and a glucosamine (GlcNAc, or GlcNS) carrying different sulfation and IdoA contents. NS domains carry high *N*-sulfation and IdoA levels; NA/NS domains carry intermediary *N*-sulfation, *N*-acetylation, and IdoA levels; NA domains are not sulfated and carry not IdoA. The GAG-CP linker biosynthetic genes are common to HSPG and CSPG biosynthetic machineries.

The biosynthesis of HSPG starts in the endoplasmic reticulum with the production of the core protein coded by a single gene (for instance *HSPG2, AGR, COLXVIIA, GPCs*, or *SDCs* that code for full time HSPG core proteins). At the Golgi, the core protein is glycosylated to first form the GAG-CP linker tetrasaccharide. This requires the expression of several genes coding for different enzymes including glycosyltransferases (*XYLT1, XYLT2, B4GALT7, B3GALT6, B3GAT3*)^4,5^, kinases (*FAM20B*)^6^, phosphatases (*PXYLP1*)^7^, and nucleosidases (*CANT1*)^8^, as well as nucleoside transporters (*SCL26A2, SLC35A2*, and *SLC35A3*)^9^ and ion transporters (*SLC10A7*)^10^ (Fig. 1).

After formation of the GAG-CP linker, HS chain initiation starts through action of EXTL3 (coded by *EXTL3*), which adds a *N*-acetyl glucosamine (GlcNAc) to the non-reducing end of the GAG-CP linker. HS chain elongation continue by sequential addition of GlcA and GlcNAc catalyzed by EXT1/EXT2 (coded by *EXT1* and *EXT2*). During chain elongation, epimerization of glucuronic acid (GlcA) into iduronic acid (IdoA) is then carried by C5 epimerase (coded by *GLCE*)^11^ and sulfation occurs at different position of the uronic acid and glucosamine residues. Sulfation of IdoA is assured by 2-*O*-sulfotransferase (*HS2ST1*), whereas sulfation of GlcNAc is assured by *N*-deacetyl-*N*-*s*ulfotransferases (coded by *NDST1, NDST2, NDST3*, or *NDST4)*, 6-*O*-sulfotransferases (coded by *HS6ST1, HS6ST2*, or *HS6ST3*), and 3-*O*-sulfotransferases (coded by *HS3ST1, HS3ST2, HS3ST3A1, HS3ST3B1, HS3ST4, HS3ST5*, or *HS3ST6*)^3,12^. During biosynthesis, the HS chain elongation can be stopped by the unproductive glycosyltransferases EXTL1 or EXTL2 (coded by *EXTL2* or *EXTL1*)^13^, whereas after secretion of the HSPG at the extracellular space 6-*O*-sulfatases SULF1 and SULF2 (coded by *SULF1* and *SULF2*) and heparanase (coded by *HPSE*) can modify the HS chain structures^14-16^. Globally, this process leads to formation of different HS structural domains^3,17^. ‘NA’ domains carry unsulfated *N*-acetylated sequences, ‘NA/NS’ domains carry HS sequences with intermediary levels of sulfation and IdoA, and ‘NS’ domains carry high levels of sulfation and IdoA^3,17^ (Fig. 1).

Regardless of their structural diversity, HS are frequently referred as a single ubiquitous molecule, possibly because their structural characterization is highly complex and yet beyond reach. Here, as an approach to reduce HSPG complexity, we propose a new strategy based in a PRISMA^18^ systematic review and analysis of well stablished pathogenic variants and emerging genetic wide association studies (GWAS) linking HSPG biosynthetic machinery gene variability to disease, susceptibility to disease, altered response to instigating stimuli, or altered behaviors. To confirm the documented clinical variants and traits in human, we compared them to the phenotypes of corresponding gene null mouse, when available. Expression patterns in different organs or tissues were retrieved from gene expression databases and information on structural features, including substrate specificities for enzymes, or protein-protein interactions for core proteins and other biosynthetic proteins, were reviewed following the PRISMA guidelines^18^. Association of this body of information resulted in a new HSPG vision that gives a better understanding of the involvement of each biosynthetic gene in the generation of HS sequences carrying structures that can play either ubiquitous or specialized roles in the maintain of tissue homeostasis, disease, behavior disorders, proper tissue response to instigating stimuli, and tissue specific vulnerability to disease.

## 2. Methods

Guidelines for Systematic Reviews and Meta-Analyses (PRISMA 2020)^18^ were followed to setup a step-wise method (Fig. 2) for selecting and filtering research items on 50 genes directly involved in the biosynthesis of HSPG (Table S1). For each HSPG biosynthetic gene, research items were organized to screen clinical variants, single nucleotide polymorphism (SNP), expression levels in different organs of the human body, and literature records databases on substrate selectivities and protein-protein interactions.

**Fig. 2.**
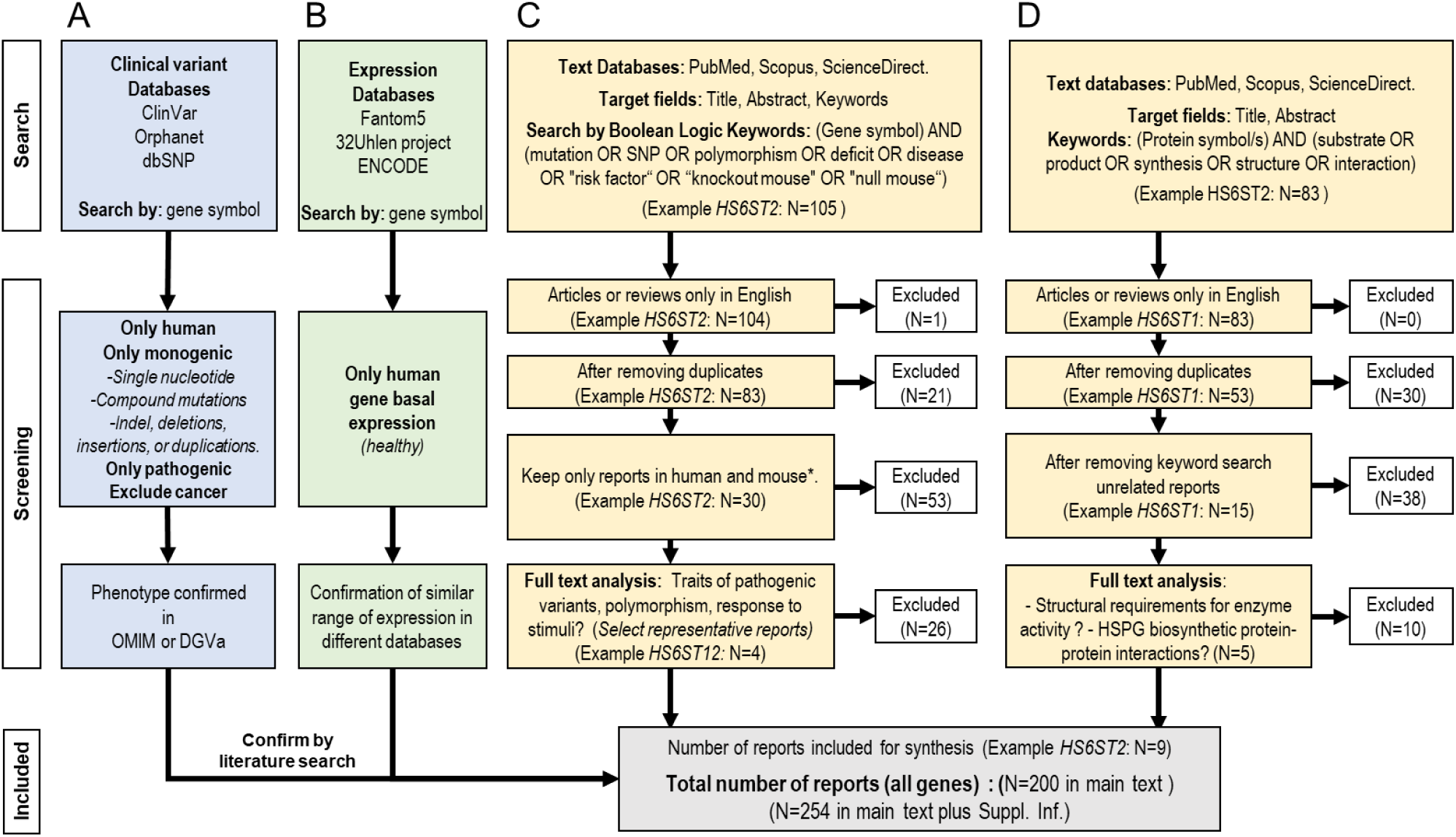
Schematic representation for collecting articles process following PRISMA^18^ recommendations. **A)** Clinical variants databases were used to document the effect of HSPG biosynthetic genes variabilities in health status. **B)** Expression databases were used to document organs or tissues in which transcript for each of the genes can be detected under basal homeostasis (healthy status). **C)** Bibliographic databases were used to document reports on the effect of genetic variability in health status and to expression in organs and tissues. **D)** Bibliographic databases were also used to document proteins interactions between biosynthetic machinery proteins and substrates selectivities for biosynthetic enzymes.

### 2.1. Search in clinical variants and SNP databases

For each human gene (Table S1), clinical variants and SNP public databases accessible without restriction were consulted by using gene names previously confirmed by MeSH (Medical Subject Headings: https://www.ncbi.nlm.nih.gov/mesh/). Data on the impact of gene variability in health status were obtained from three main databases (Fig. 2A): 1) the ClinVar archive (database ClinVar: https://www.ncbi.nlm.nih.gov/clinvar/intro/), which gives information on correlations between genetic variations and overt phenotypes or health status with history of interpretations, 2) the Orphanet archive (Database Orphanet: https://www.orpha.net/), which considers clinical presentation based on published scientific articles and expert reviews, and 3) the dbSNP archive (database: https://www.ncbi.nlm.nih.gov/snp/), which is a free public archive for genetic variation within and across different species. For clinical variants, only monogenic variants (indel, deletions, duplications, insertions, and single nucleotide) were considered. Clinical significance was further confirmed in web platforms including Online Mendelian Inheritance in Man (OMIM) (https://www.omim.org/) and the Database of Genomic Variants Archive (DGVa database: https://www.ebi.ac.uk/dgva/). Information on clinical variants overt phenotypes in human were confirmed by literature research on case reports and on reports on the corresponding gene null mice (Methods section 2.3 and Fig. 2C-2D).

### 2.2. Search in expression databases

Information on the basal expression of the HSPG biosynthetic genes (Table S1) in different tissues or organs of the healthy adult human body was retrieved from open access transcriptomic data bases including: 1) the RNA-seq 32Uhlen project (database 32Uhlen: http://www.proteinatlas.org/humanproteome), which analyzed 32 different tissues from 122 human individuals, 2) the RNA-Seq CAGE (Cap Analysis of Gene Expression) in the RIKEN FANTOM5 project (database FANTOM5: http://fantom.gsc.riken.jp/data/), which analyzed several healthy adult human tissues, and 3) the ENCODE strand-specific RNA-seq of 13 human tissues from Michael Snyder’s lab (Database ENCODE: https://www.encodeproject.org/). All these RNA-seq databases are considered in the expression atlas (http://www.ebi.ac.uk/gxa)^19^, as they use standardized methods that allow the determination of baseline gene expression across controlled biological replicates checked for high quality (good quality raw files and good quality reference genome build are available). The original raw and processed data files can be found in the ArrayExpress platform (https://www.ebi.ac.uk/arrayexpress/), in which the raw single-channel microarray intensities were normalized either using an oligo package from Bioconductor (Affymetrix data) or by quantile normalization by the limma package (Agilent data). The data quality was assessed using the arrayQualityMetrics package in R. Quality-trimmed reads were mapped to the latest version of the reference genome from Ensembl using TopHat2. For each HSPG biosynthetic gene, the number of mapped reads per gene were summed (raw counts) using htseq-count and represented as average transcripts per million (TPM). When transcripts were detected in a tissue, regardless of their level of expression, a gene was considered to be basally expressed in that tissue. To illustrate gene expression, the best TPM value from the different databases was included in the analysis.

### 2.3. Search in literature databases

PRISMA guidelines^18^ were used to carry two campaigns of literature research for each gene. The first campaign was performed to confirm information obtained from clinical variants databases (section 2.1 and Fig. 2A) by search of clinical variants case reports and reports on the corresponding gene null mouse (Fig. 2C). The second campaign was performed to retrieve information on each gene product functionally, including structural features for substrate specificity and selectivity for enzymes, and on protein-protein interactions for core proteins and other biosynthetic proteins (Fig. 2D). Three literature search database thesauruses (PubMed, Scopus, and Science Direct) were employed for a systematic search facilitating an extensive inclusion (Fig. 2C and 2D). For each search engine, 2 subsets of pool keywords and Boolean logic were conducted for each of the 45 genes. The first keywords subset ((‘gene symbol’) AND (mutation OR SNP OR polymorphism OR deficit OR disease OR “risk factor” OR “knockout mouse” OR “null mouse”)) was used to review the effect of genetic variability in health status (Fig. 2C). The second keywords and Boolean logic subset was used to review the substrates specificities for enzymes or protein-protein interactions ((‘protein name’ OR ‘protein symbol’) AND (substrate OR “substrate structure” OR “substrate specificity” OR interaction OR “interaction specificity”)) (Fig. 2D).References were screened in accordance with title and abstract. Only research articles and literature reviews were considered. Only reports in English language were included. We did not specify a time frame in our literature search. An email alarm system, on each database, was created for notifications on new publications related to the search terms. All reports sorted in the three databases were exported to a common file for each gene in a bibliographic management system (EndNote) and an automatic deduplication tool was applied to discard duplicated reports. Then, reports in species other than human and mouse and reports in cancer cell lines were systematically excluded, unless any example of SNP could be found in human. A pilot user study was then conducted for examining full text reports and representative references were selected and pooled in a single bibliographic management file using EndNote. The reference list of the retrieved reports was also explored, articles non available were requested to authors via email, otherwise references could not be included. Finally, our systematic review included a synthesis of 254 peer-reviewed reports that were checked and review as representative of the full retrieved data (200 reports cited in main text and additional 54 only cited in Suppl. Inf.).

## 3. Results

### 3.1. Essential and non-essential HSPG biosynthetic genes

A first clustering of the HSPG biosynthetic genes was performed on the bases of their requirement for normal embryonic development and homeostasis, as reported by clinical variants databases. Information on gene essentialness were confirmed by literature records in cases reports and/or from reports on the corresponding gene null mouse (see extended review on genes in Suppl. Inf.). Genes considered essential for normal embryonic development and homeostasis were those for which genetic variability in human is causative of diseases or syndromes, and which corresponding null mouse dies or shows overt phenotypes, consistent with that observed in clinical variants. Genes for which genetic variability has not been reported as causative of human diseases or syndromes, and/or for which the corresponding null mouse is viable, fertile, lives normally, and shows non-overt phenotypes in the absence of stimuli, were considered non-essential for normal embryonic development and homeostasis (Suppl. Inf.). However, most of the ‘Non-essential’ genes show some genetic variability associations with altered responsiveness to stimuli, altered behaviors, or are involved in vulnerability to altered conditions health conditions or behaviors (Suppl. Inf.). Based in these criteria, 2 clusters of genes were formed, the first cluster is referred as ‘Essential’, and the second as ‘Non-essential’ (Fig. 3). To avoid loss of clustering, HSPG biosynthetic genes involved in tumor development and growth was not considered, as most HSPG are altered in cancer^16,20,21^.

**Fig. 3.**
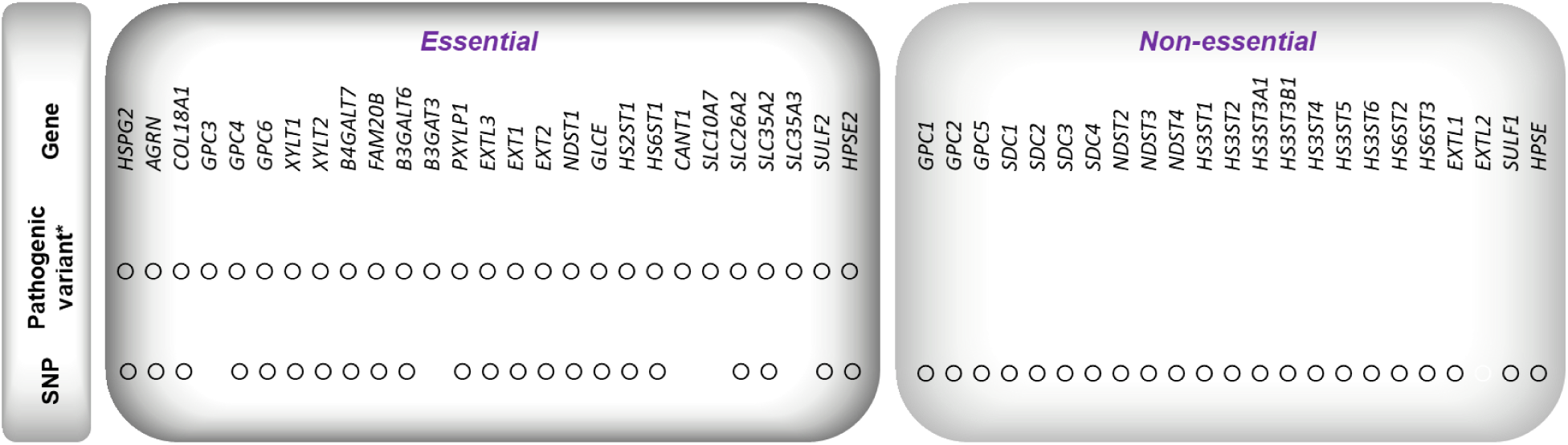
First clustering of HSPG genes depending on their requirement for normal development and homeostasis. ‘Essential’ genes are those for which genetic variability in human is causative of diseases or syndromes and/or the corresponding null mouse dies or shows overt phenotypes consistent with what is observed in clinical variants. *They can show SNP. ‘Non-essential’ genes are those for which genetic variability has not been reported as causative of human diseases or syndromes and/or for which the corresponding null mouse is viable, fertile, lives normally, and shows non-overt phenotypes in the absence of stimuli, although many of them show SNP associations with altered responsiveness to stimuli, altered behaviors, or are involved in vulnerability to disease.

### 3.2. Ubiquitous and tissue restricted HSPG biosynthetic machinery genes

To get additional information on Essential and Non-essential HSPG biosynthetic genes in relation with their basal expression in human, genes were re-clustered depending on their expression in healthy organs or tissues, as documented in RNAseq databases (see Methods). Genes which transcripts were detectable in all analyzed organs were considered as ‘widely expressed’, whereas genes which expression was only detected in some tissues/organs were considered as ‘restrictedly expressed’ (Fig. 4 and Supp. Inf.). This transcription-based re-clustering resulted in the HSPG biosynthetic genes organization in four groups of genes (Fig. 4): Group I, Group III, Group III, and Group IV, each of which is described below (simplified clustering is presented in Fig. 5). A review on each gene is detailed in Suppl. Inf.

**Fig. 4.**
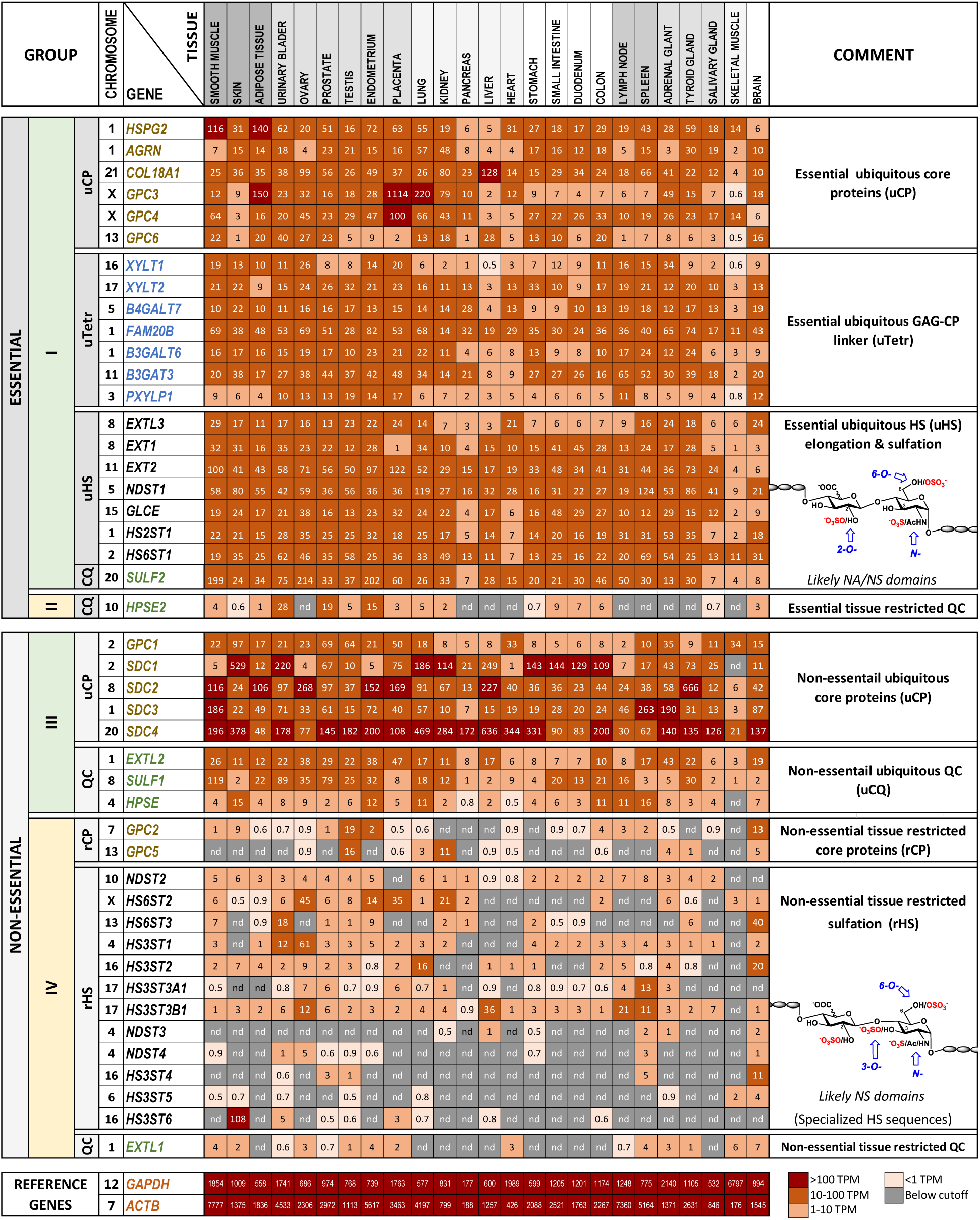
The human HSPG biosynthetic machinery genes organized by gene essentiality and expression levels. HSPG biosynthetic genes were first separated in ‘Essential’ or ‘Non-essential’ for normal development and homeostasis depending on their requirement for normal human development and homeostasis and/or the phenotype of the corresponding gene null mouse (Suppl. Inf). Then, genes were re-clustered depending on their expression (transcript par million TPM) in a variety of human organs or tissues, as documented from RNAseq databases (FANTOM5: http://fantom.gsc.riken.jp/data/; ENCODE: https://www.encodeproject.org/; and 32Uhlen: http://www.proteinatlas.org/humanproteome/tissue+specific). Genes were organized in 4 main groups depending on their basal expression levels in adult human. CQ stands for ‘Quality Control’ that can arrest HS elongation during synthesis or remodel HS structure after secretion to the extracellular space. Ion and nucleotide transporters coding genes are not included.

**Fig. 5.**
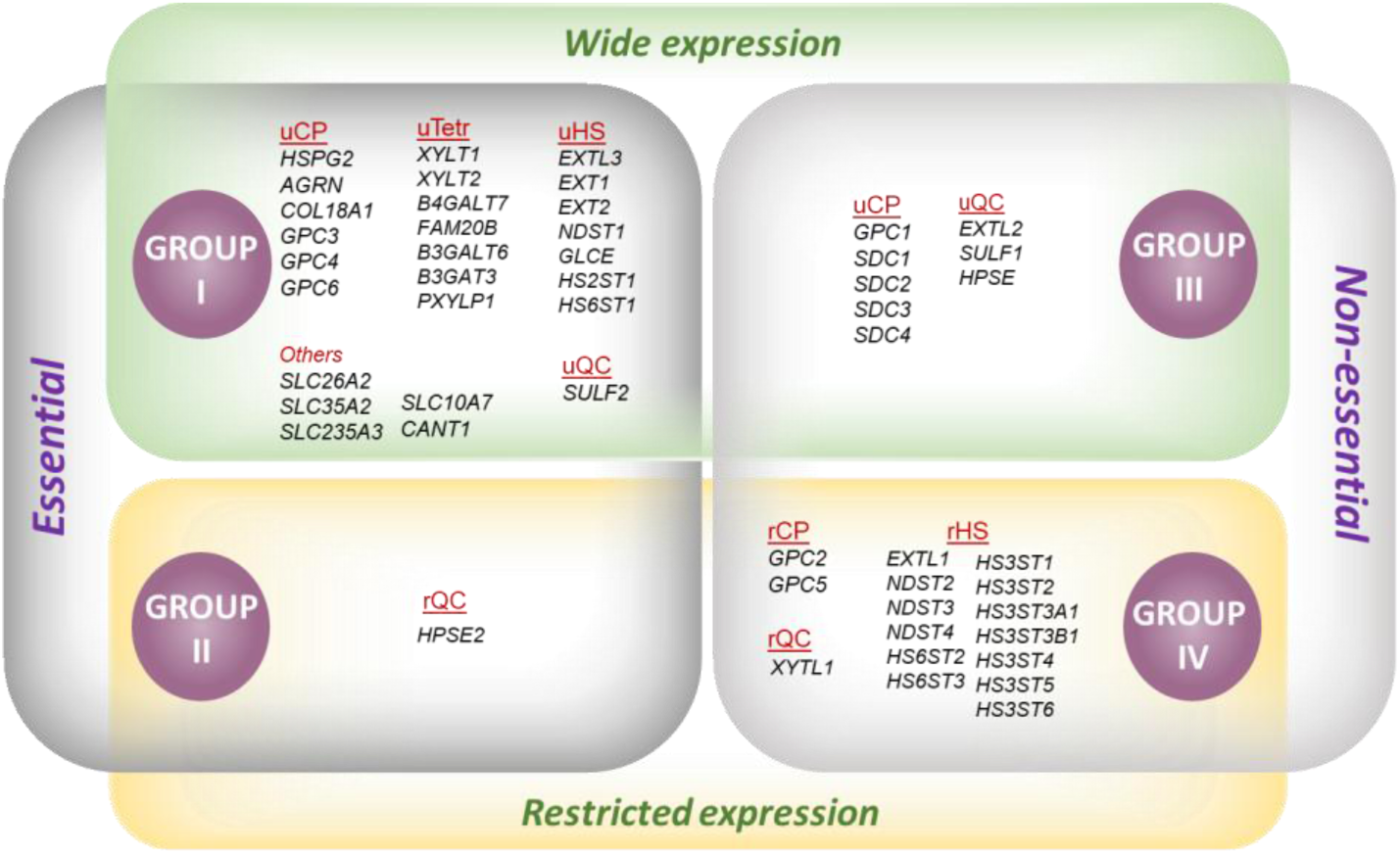
Simplified organization of the HSPG biosynthetic machinery genes. After a first clustering in ‘Essential’ or ‘Non-Essential’, GSPG biosynthetic genes were clustered depending on whether they show restricted expression or wide expression in human tissues under basal physiologic conditions. This dual organization allowed formation of four groups of genes: **Group I** integrates Essential and widely expressed genes coding for ubiquitous HSPG core proteins (uCP), ubiquitous GAG-CP linker tetrasaccharide (uTetr), and ubiquitous HS sulfate (uHS) biosynthetic enzymes and transporters. **Group II** integrates an Essential and tissue-restricted gene involved in the control of post-synthetic HS modifications (indicated as rCQ, for tissue restricted quality control). **Group III** integrates ‘Non-essential widely expressed genes’ coding for ubiquitous HSPG core proteins (uCP) and widely expressed genes involved in HS post-synthetic modifications (uQC). **Groupe IV** integrates ‘Non-essential restrictedly expressed genes’ coding for tissue restricted core proteins (rCP), tissue restricted HS (rHS) biosynthetic enzymes, and tissue restricted genes involved in post-synthetic modifications (indicated as rCQ, for tissue restricted quality control).

#### 3.2.1. Group I - Essential HSPG biosynthetic genes that are ubiquitously expressed

The Group I of HSPG biosynthetic genes clustered widely expressed genes that are essential for normal development and homeostasis (Fig. 4, simplified Fig. 5, and Suppl. Inf). This included genes coding for pericellular core proteins *HSPG2*^22-25^, *AGRN*^26-29^, and *COL18A1*)^30-32^, for some membrane-associated core proteins (*GPC3*^33-35^, *GPC4*^36,37^, *GPC6*^*38-40*^), and for all the genes coding for the GAG-CP linker biosynthetic proteins, including glycosyl transferases (*XYLT1*^41,42^, *XYLT2*^43^, *B4GALT7*^44,45^, *B3GALT6*^46,47^, and *B3GAT3*^48^), kinase FAM20B (*FAM20B*)^49^, phosphatase (*PXYLP1*)^7^, ion or nucleotide transporters (*SLC10A7*^10^, *SLC26A2*^*50*^, *SCL35A2*^*51*^, and *SCL35A3*^9,52^), or other Golgian enzymes (*CANT1*)^8^. Importantly, Group I also clustered genes coding for the glycosyltransferases that assure HS chain initiation and elongation (*EXTL3*^53,54^, *EXT1*^55^, *EXT2*^56^), for epimerase (*GLCE*)^57^, and for three sulfotransferases (*NDST1, HS2ST1*, and *HS6ST1*)^58-61^. Moreover, *SULF2*, which codes for one of the extracellular 6-*O*-sulfatase that generates biologically active HS sequences resulting from the cleavage of 6-*O*-sulfated groups^14,62^, also clustered in Group I. Altogether, the enzymes coded by the ubiquitous HS chain biosynthetic genes can then produce ubiquitous HS sequences (uHS), which fine structures might depend on the expression levels of each biosynthetic gene in each particular cell or tissue^63-65^, as well as in their transcriptional regulation and translational control^66^. Together, this body of data suggests that the Group I HSPG biosynthetic machinery can, by itself, produce HSPG carrying biologically relevant HS sequences or chains that are essential for normal development and homeostasis.

#### 3.2.2. Group II - Essential tissue restricted HSPG biosynthetic genes

Group II clustered HSPG biosynthetic genes that, although essential, show tissue restricted expression. Based in current available data and literature records, only *HPSE2* clustered in this group (Fig. 4, Fig. 5, and Suppl. Inf). *HPSE2* codes for the enzymatically inactive heparanase 2. Although it does not affect HS structures, heparanase 2 can bind to these polysaccharides with higher affinity than the enzymatically active heparanase (Group III), avoiding its enzymatic action and maintaining long HS chains in their biological environment^67,68^. *HPSE2* is here reported as the only gene in Group II, but the evolving literature and clinical reports might further enrich the number of genes in this group.

#### 3.2.3. Group III - Non-essential HSPG biosynthetic genes that are ubiquitously expressed

Group III clustered HSPG biosynthetic genes that although widely expressed are non-essential for normal development and homeostasis (Fig. 4, Fig. 5, and Suppl. Inf.). However, genes in Group III appeared to be required for appropriate tissue responsiveness to stimuli and, for several of them, SNP is associated with increased or decreased vulnerability to disease, or to altered conditions or behaviors (Supp. Inf). Interestingly, all syndecans core proteins coding genes (*SDC1*^*69-71*^, *SDC2*^72^, *SDC3*^73-76^, *and SDC4*^*77-79*^) and one glypican (*GPC1*^80,81^) clustered in this group (Fig. 4). Concerning the HS chain biosynthetic genes, none of these clustered in this group. Instead, Group III clustered genes involved in the control of HS levels and quality, including *EXTL2*^82,83^, *HPSE*^*84-86*^ and *SULF1*^70,87-89^ (Fig. 5). Globally, data from GWAS, knockout mice, and literature research indicate that genes in Group III are not essential for normal development and homeostasis but are required for proper response to stimuli and or involved in vulnerability or resistance to disease and altered behaviors.

#### 3.2.4. Group IV - Non-essential HSPG biosynthetic genes that are restrictedly expressed

Group IV of HSPG biosynthetic machinery genes clustered all genes that are expressed in a tissue restricted manner (Fig. 4) and which genetic variability is not causative of lethality or disease, although SNP might be associated to resistance or vulnerability to pathologic conditions, to altered response to stimuli, and to altered behaviors (Fig. 4, Fig. 5, and Suppl. Inf). Genes in Group IV included those coding for two core proteins (*GPC2*^90-92^ and *GPC5*^93-95^), three NDSTs (*NDST2*^96-98^, *NDST3*^99-101^, *and NDST4*^102,103^), two HS6STs (*HS6ST2*^104,105^, *HS6ST3*^106,107^), all HS3STs (*HS3ST1*^108-110^, *HS3ST2*^111,112^, *HS3ST3A1*^113-115^, *HS3ST3B1*^115^, *HS3ST4*^116^, *HS3ST5*^117^, and *HS3ST6*^118,119^), and one HS levels regulating genes (EXTL1^120^). Based in current available data and literature records, it appeared that, at least in the absence of instigating stimuli, these genes are non-causative of disease and are not essential for normal development and homeostasis (Supp. Inf). Accordingly, their corresponding gene null mice, when available, develop normally, are fertile, and show non deleterious phenotypes (Supp. Inf). Depending on the gene, mice can show inappropriate response to instigating stimuli during processes including wound healing, angiogenesis, inflammation, immunity, infection, and response to neurological stimuli leading to altered adaptative behaviors (as alertness and satiety during feeding, and response to stress) (Suppl. Inf.). Moreover, SNP in Group IV genes, along or in combination with SNP in other loci, have been associated to vulnerability or resistance to diseases, to infection, and to impaired angiogenesis, inflammation, immunological response, altered behaviors, etc. (Fig. 3 and Suppl. Inf.). It is to note that all the HS sulfotransferases coding genes that did not cluster in Group I clustered in this group, including *NDST2, HS6ST2, HS6ST3*, and all *HS3STs*. Thus, based in information from knockout mice, GWAS data, and literature research, it appeared that the tissue restricted rHS sequences are those required for appropriate adaptive behaviors and responsiveness to stimuli in tissues in which each gene is specifically expressed.

### 3.3. Functional aspects of the HSPG core proteins

#### 3.3.1. Essential pericellular core proteins

Pericellular HSPG are essential constituents of basement membranes, the highly specialized extracellular matrices that tie the epithelium to connective, endothelial, muscular, neuronal, and other tissues^121^. The main pericellular HSPG core proteins are perlecan, agrin, and collagen 18, respectively coded by *HSPG2, AGRN*, and *COL18A1*, which clustered in Group I (Table 1, Fig. 5, and Suppl. Inf.). Perlecan is mainly expressed in skeletal, musculoskeletal, and cardiovascular basement membranes^23,122^. Hence, *HSPG2* variants affect these tissues with severities that depend on the variant, as observed in Stuve-Wiedemann, Schwartz–Jampel and lethal Kniest-Like syndromes^23,25^, which traits are recapitulated in the *Hspg2*^*-/-*^ mouse^22^. Similarly, clinical variants in *AGRN*, which codes for the agrin core protein characteristically expressed in neuromuscular junctions^122^, result in a myasthenic syndrome, recapitulated in the *Agrn*^-/-^ mouse^27,29^. Collagen XVIII additionally supports basement membranes in some tissues^32^, as suggested by the traits observed in *COL18A1* variants that lead to eye defects, occipital encephalocele (Knobloch syndrome), and other overt phenotypes recapitulated in the *Col18a1*^-/-^ mouse^30,31^. In addition to the importance of pathogenic variants, SNP has been associated with risk to altered health condition or disease (Table 1, Fig. 3). For instance, *HSPG2* SNP has been associated with tardive dyskinesia^123^ whereas *COL18A1* SNP has been associated with pelvic organ prolapse^124^. Globally, these data confirm the essentialness of ubiquitous pericellular HSPG core proteins, and the importance of their likely-ubiquitous HS chains, in the pericellular matrix organization^1^.

**Table 1.**
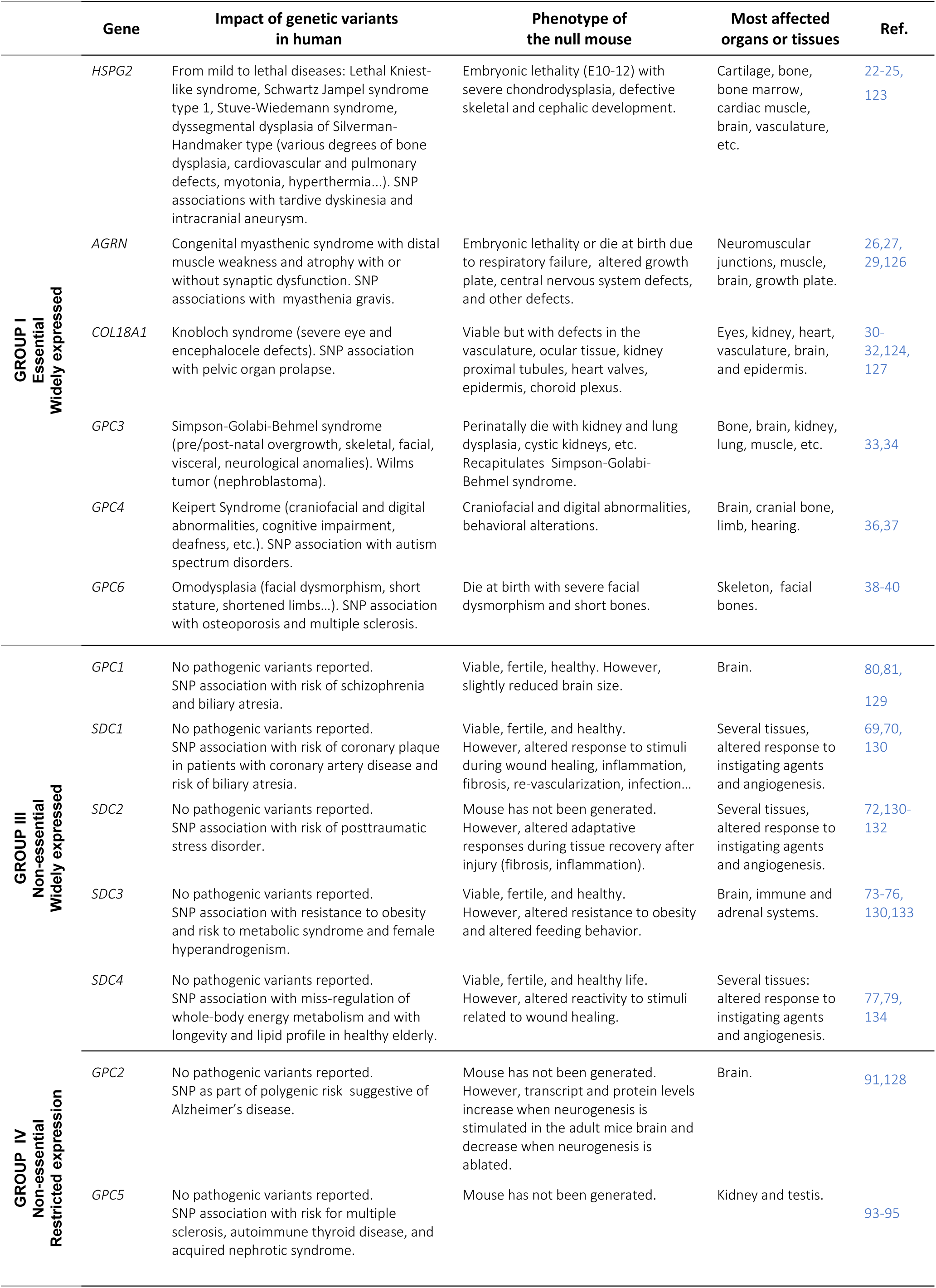
Genes coding for ubiquitous and tissue restricted HSPG core proteins. Genes are organized based on their expression in human tissues and traits of their clinical variants and/or phenotype of null mice.

#### 3.3.2. Essential cell membrane-associated core proteins

Among HSPG membrane-associated core proteins coding genes, only *GPC3, GPC4*, and *GPC6* clustered in Group I (Table 1, Fig. 5, and Supp. Inf.). By keeping the HS chains very close to cell membranes, glypicans are known to bring HS binding proteins near to their high affinity receptors^125^. Proteins requiring this proximity includes fibroblast growth factors, bone morphogenetic proteins, wints, hedgehogs, and several other proteins involved in cell signaling^125^. Among *GPCs* pathogenic variants, those in *GPC3* are the most severe, as shown by pre-and post-natal overgrowth with skeletal, visceral, and neurological abnormalities observed in the X-linked Simpson-Golabi-Behmel syndrome resulting from *GPC3* variability^34^ and recapitulated in the *Gpc3*^*-/-*^ mice^33^. Similar traits are observed in *GPC4* variants responsible of Keipert syndrome^36^, characterized by craniofacial and digital abnormalities, deafness, and synaptic impairment, and in *GPC6* variants responsible of omodysplasia-1^37^ with facial dysmorphism, short stature, and shortened limbs, recapitulated in the knockout mice^39^. In addition to pathogenic genetic variability, SNP in *GPC4* and *GPC6* has been associated to risk to altered conditions (Table 1 and Suppl. Inf). These data confirm the essentialness of the ubiquitous *GPC3, GPC4, GPC6* in normal development and homeostasis and suggest that glypican, at least glypicans 3, 4 and 6, are the main HSPG that carry the HS involved in essential cell signaling. Whether these HS are made by the also ubiquitous HS biosynthetic genes is unknown.

#### 3.3.3. Non-essential cell membrane-associated HSPG core proteins

Excepting the essential *GPC3, GPC4, GPC6*, all other cell membrane-associated core proteins coding genes (*GPC1, GPC2, GPC5, SDC1, SDC2, SDC3*, and *SDC4*) clustered in Groups III and IV, both characterized for being non-essential for normal development but required for appropriated response to stimuli, as during wound healing, angiogenesis, inflammation, immune response, normal behaviors, etc. (Table 1, Fig. 5, and Supp. Inf.). For instance, SNP in *GPC1* loci was associated to biliary atresia^80^ and schizophrenia^81^, whereas *GPC2*, expressed in brain (Fig. 4) was involved in proper response to neurogenic stimuli^91^. Similarly, the kidney, testis, and adrenal and thyroid glands predominant *GPC5* (Fig. 4) was involved in podocyte response to injury and nephropathy progression in type 2 diabetes patients^135^ and SNP was associated with acquired nephrotic syndrome, autoimmune thyroid disease, and risk for multiple sclerosis^93-95^. Similarly, syndecans are required for appropriate tissue response to inflammation, infection, wound healing, angiogenesis, neurological stimuli, etc.^136-138^. In agreement with the role of *SCD* genes in appropriate response to stimuli, the *Sdc1*^-/-^ mouse showed altered capacity to recover from microbial infection or ischemia^139^ and the *Sdc4*^-/-^ mouse showed delayed wound healing and impaired angiogenesis^134^. Although the *Sdc2*^*-/-*^ mouse has not yet been generated, syndecan 2 has been involved in fibrosis and inflammation^131,132^. Moreover, SNPs in some of these genes have been associated with risk to develop diseases involving altered adaptative behaviors. For instance, *SDC1* polymorphism has been associated with coronary plaque formation in coronary artery disease patients^69^, with multiple sclerosis^70^, and with renal malfunction in adolescents with excess weight^71^. *SDC2* polymorphism has been associated to posttraumatic stress disorders in post war veterans^72^. *SDC3* polymorphism has been associate to satiety dysregulation and obesity^74,76^, metabolic syndromes^73^, and schizophrenia^75^. *SDC4* polymorphism was associated with risk of metabolic syndromes and obesity^77^ and with longevity in healthy elderly Italian^79^. Together, these data indicate that Group III and IV core proteins, and possibly their HS chains, are required for appropriate responsiveness to stimuli and adaptative behaviors rather than for the regulation of essential physiological processes.

### 3.4. The GAG-CP-linker tetrasaccharide pathway first assures HSPG over CSPG biosynthesis

After synthesis of the core protein, the GAG-CP linker tetrasaccharide (Fig. 1) is synthetized by an ubiquitous biosynthetic machinery involving several genes which, with exception of XYLT1 and XYLT2^3^, have not paralogue genes able to assure compensation. Because this machinery is common to HSPG and CSPG, it can then be expected that defects in any of the ubiquitous GAG-CP linker biosynthetic genes will affect production of both proteoglycans in all tissues. However, a skeletal phenotype predominantly affecting CS is mainly observed in linkeropathies, the family of diseases caused by variability in GAG-CP linker biosynthetic genes^140^. Here, the integrative analysis of clinical variants, gene expression, enzymes selectivities, and protein clusters data (Table 2, Fig. 4, and Supp. Inf.) indicates that the GAG-CP linker biosynthetic pathway is organized to preferentially assure production of HSPG over that of CSPG (Fig. 6A), as detailed below.

**Table 2.** Ubiquitous GAG-CP linker tetrasaccharide biosynthetic genes. Genes are organized based on their expression in human tissues and traits of their clinical variants and/or phenotype of null mice.

| Gene | Impact of genetic variants in human | Phenotype of the null mouse | Most affected organs or tissues | Ref. |
| --- | --- | --- | --- | --- |
| XYLT1 | Desbuquois dysplasia (DBQD) type 2 (also known as Baratela–Scott syndrome), characterized by with short stature, joint laxity, advanced carpal ossification, mental retardation. | Non-available. However, a mouse carrying a *Xylt1* mutation shows chondrocyte premature maturation, early ossification, and dwarfism. | Connective and skeletal tissue, brain. | 41,42 |
| XYLT2 | Spondyloocular syndrome (post-developmental osteoporosis, cataract, hearing and learning impairment). | Develops normally but show post-developmental defects in liver, biliary epithelial and renal cysts, low body weight, adipose tissue loss and lipodystrophy. | Skeletal, adipose, and connective tissues, liver, eye, bone, brain, kidney. | 43,135, 147 |
| B4GALT7 | Spondylodysplastic Ehlers-Danlos Syndrome (spEDS)-progeroid type (connective and skeletal tissues with radioulnar synostosis) and Reunion Island Larsen-like syndrome (large spectrum). | Not reported. | Bone, cartilage, skin, brain, eye. | 44,45, 148 |
| FAM20B | Short limb dysplasia resembling XYLT1-related DBQD with mid-face and thoracic hypoplasia (leading to respiratory failure), very short stature, mesomelic limbs shortening and multiple joint dislocations. | Dies as embryo and shows severely stunted growth, multisystem organ hypoplasia, and delayed development of the skeletal system, eyes, lung, gastrointestinal tract, and liver. | Multiorgan hypoplasia, delayed growth and development. | 49,149, 150 |
| B3GALT6 | spEDS progeroid type 2 (affecting connective and skeletal systems with sever skeletal affections), spondyloepimetaphyseal dysplasia with joint laxity, learning defects. | Not reported. | Bone, cartilage, skin tendons, ligaments, brain. | 46,47 |
| B3GAT3 | Larsen-like syndrome with variable phenotypes similar to Antley-Bixler, Shprintzen-Goldberg, and Geroderma osteodysplastica syndromes. | Embryonic lethality before the 8-cell stage because failed cytokinesis. | Bone, cartilage, skin tendons, ligaments, heart, brain. | 48 |
| CANT1 | Various skeletal phenotypes related to DBQD and Larsen syndrome with growth retardation, short extremities, progressive scoliosis, joint laxity, severe prenatal and postnatal growth retardation, etc. | Shows the continuum of skeletal dysplasia phenotypes including Desbuquois dysplasia and multiple epiphyseal dysplasia. | Skeletal and connective tissues, brain. | 8,151 |
| SLC10A7 | Shortened long bones, growth plate and tooth enamel anomalies. | Short stature, amelogenesis imperfecta, and skeletal dysplasia with scoliosis. | Skeletal and connective tissues, brain. | 10 |
| PXYLP1 | No pathogenic variants reported possibly because lethal. SNP association with good prognosis in cancer immunotherapies. | Not reported. | Unknown | 152 |

#### 3.4.1. The GAG-CP linker biosynthetic machinery

Biosynthesis of GAG-CP linker is initiated by addition of a xylose (Xyl) from UDP-Xyl to specific serine residues in core proteins (Ser-CP)^2,3^. This is assured by XYLT1 during development and by both XYLT1 and XYLT2 after bird^42^ (Fig. 6A and Supp. Inf.). During development, *XYLT1* is highly expressed by chondroblasts^42^, the cells that secrete a CSPG aggrecan rich matrix that forms the cartilage template upon which bone is formed^141^. Indeed, as endochondral ossification of the cartilage elements ensues, the expression of aggrecan by differentiating chondrocytes predominates over expression of other proteoglycan core proteins^142^. Thus, to assure all proteoglycans glycosylation in this high aggrecan content, XYLT1 binds to aggrecan core protein with lower affinity than to other core proteins^41,143^, including to the small leucine rich (SLRP) CSPG decorin that stabilizes collagen fibrils in soft tissues, as skin and cornea^144^. The higher affinity of XYLT1 for decorin over aggrecan splits the GAG-CP linker biosynthetic pathway in a CSPG-decorin path and a HSPG/CSPG-aggrecan path (Fig. 6A), in which rich aggrecan synthesis is assured by an abundant substrate pool. Thus, during development, a defective *XYLT1* should primarily assure the decorin path, principally affecting the CSPG-aggrecan path. Accordingly, *XYLT1* variability results in Desbuquois dysplasia type 2 (known as Baratela–Scott syndrome)^41^, characterized by severe skeletal growth retardation with multiple dislocations, joint laxity, advanced carpal ossification, with no skin overt phenotypes, indicating that the CSPG-decorin path is unaffected (Table 2 and Supp. Inf.). After birth, under physiologic conditions, *XYLT1* expression decreases and *XYLT2* is expressed. XYLT2 shows similar affinity for both decorin and aggrecan core proteins, possibly because after birth the abundant aggrecan synthesis is not any more required and HSPG and CSPG syntheses need to be balanced. Although at different extents depending on the tissue, both XYLT1 and XYLT2 are expressed after birth (Fig. 4), Thus, defects in *XYLT2* should not affect development and after birth they might be compensated by *XYLT1*, at least in some extent and in the tissues in which the enzyme is expressed. Accordingly, during development *XYLT2* variants show no clinical significance, but after birth they cause the post-developmental spondyloocular disorder characterized by osteoporosis, cataract, hearing, and learning impairment^43,135^.

On the continuation of the GAG-CP linker biosynthesis, B4GALT7 adds galactose (Gal) to Xyl-(Ser-CP)^145^ (Fig. 6A). B4GALT7 shows similar affinity for aggrecan and decorin core proteins, indicating that defective B4GALT7 should affect both CSPG-decorin and HSPG/CSPG-aggrecan paths. Accordingly, *B4GALT7* variants lead to B4GALT7-related Ehlers-Danlos syndrome (spondylodysplastic EDS type 1)^45^, which traits are like those caused by *XYLT1* mutations (CSPG-aggrecan path affected) with additional soft tissues overt phenotypes including hyperextensible, soft, thin, translucent, and doughy skin (CSPG-decorin path is affected) (Table 2 and Supp. Inf.)^45^.

In the HSPG/CSPG-aggrecan path, the resultant Gal-Xyl-(Ser-CP) is then phosphorylated by FAM20B to form Gal-Xyl(P)-(Ser-CP) (Fig. 6A), which is the preferred B3GALT6 substrate that additionally promotes the HSPG/CSPG-aggrecan path. Indeed, in further steps of the GAG-CP linker synthesis, phosphorylated products will become the preferred substrates that nourish the HSPG path^49,146^. This is consistent with the high lethality observed in the rare *FAM20B* clinical variants, which show a phenotype like that observed in *XYLT1*-Desbuquois dysplasia (CSPG-aggrecan path is affected) but additionally affecting many other organs^49^ (HSPG path is affected) (Fig. 6A). In the next step of the GAG-CP synthesis, B3GALT6 transfers a second Gal to Gal-Xyl(P)-(Ser-CP) in the HSPG/CSPG-aggrecan path and to Gal-Xyl-(Ser-CP) in the CSPG-decorin path^145^, nourishing both pathways (Fig. 6). Again, the phosphorylated product is the preferred B3GAT3 substrate that will then favor the HSPG path over the CSPG-aggrecan and CSPG-decorin paths (see below). Accordingly, *B3GALT6* mutations lead to B3GALT6-related spondylodysplastic Ehlers-Danlos syndrome (spEDS), characterized by bone dysplasia, joint laxity (CSPG-aggrecan path is affected), and mild skin hyper elasticity^47^ (CSPG-decorin path is affected) (Table 2). Subsequently, depending on substrate pool levels, B3GAT3 will act alone or in association with PXYLP1, for which it shows a high affinity. Indeed, when complexed to PXYLP1, for which B3GAT3 shows the high affinity, GlcA addition occurs followed by immediate Xyl dephosphorylation to form GlcA-Gal-Gal-Xyl-(Ser-CP), which is the preferred substrate for EXTL3 that starts HS chain initiation^7,146,153^ (Fig. 6A and 6B). Abundance of GlcA-Gal-Gal-Xyl-(Ser-CP) substrate pool allows CSGALNACT1 action on substrates that did not integrate the HS path. Moreover, in associating with B3GAT3 un-complexed PXYLP1, CSGALNACT1 can also act in Gal-Gal-Xyl(P)-(Ser-CP), which is the preferred substrate for CSGALNACT1, starting CS chain initiation^7,146,153^. Thus, higher expression levels of B3GAT3 compared to PXYLP1 (Fig. 4) and high affinity between PXYLP1 and B3GAT3 can favor equilibrium through the HS synthesis. PXYLP1 un-complexed B3GAT3 (Fig. 6A). Therefore, in defective *EXTL3*, its substrate excess allows the initiation of CS chain synthesis by CSGALNACT1 along or in association with PXYLP1^146,153^, favoring aggrecan synthesis (Fig. 6A and 6B)^7,146,153^. Thus, the phosphorylated and non-phosphorylated substrate pools and the levels of expression of B3GAT3, PXYLP1, XYLT3, and CSGALNACT1, at least, will participate to the selection of the proteoglycan that will be preferentially elongated^146^. Globally, based in literature data, clinical variants and expression databases, our analysis indicate that the CSPG and HSPG overlapping paths can concomitantly occur with crossed pathways, which preferences are driven by abundancy of substrates pools, substrates selectivities, levels of expression, and protein-protein interactions. In case of defective genes, the preferred maintain of the HSPG path is supported by the mixed skeletal and soft tissue phenotypes observed in *B3GAT3* clinical variants responsible of Larsen, Antley-Bixler, Shprintzen-Goldberg, Geroderma osteodysplastica, and spEDS^48^ and other linkeropathies (Tables 2 and 3).

**Fig. 6.**
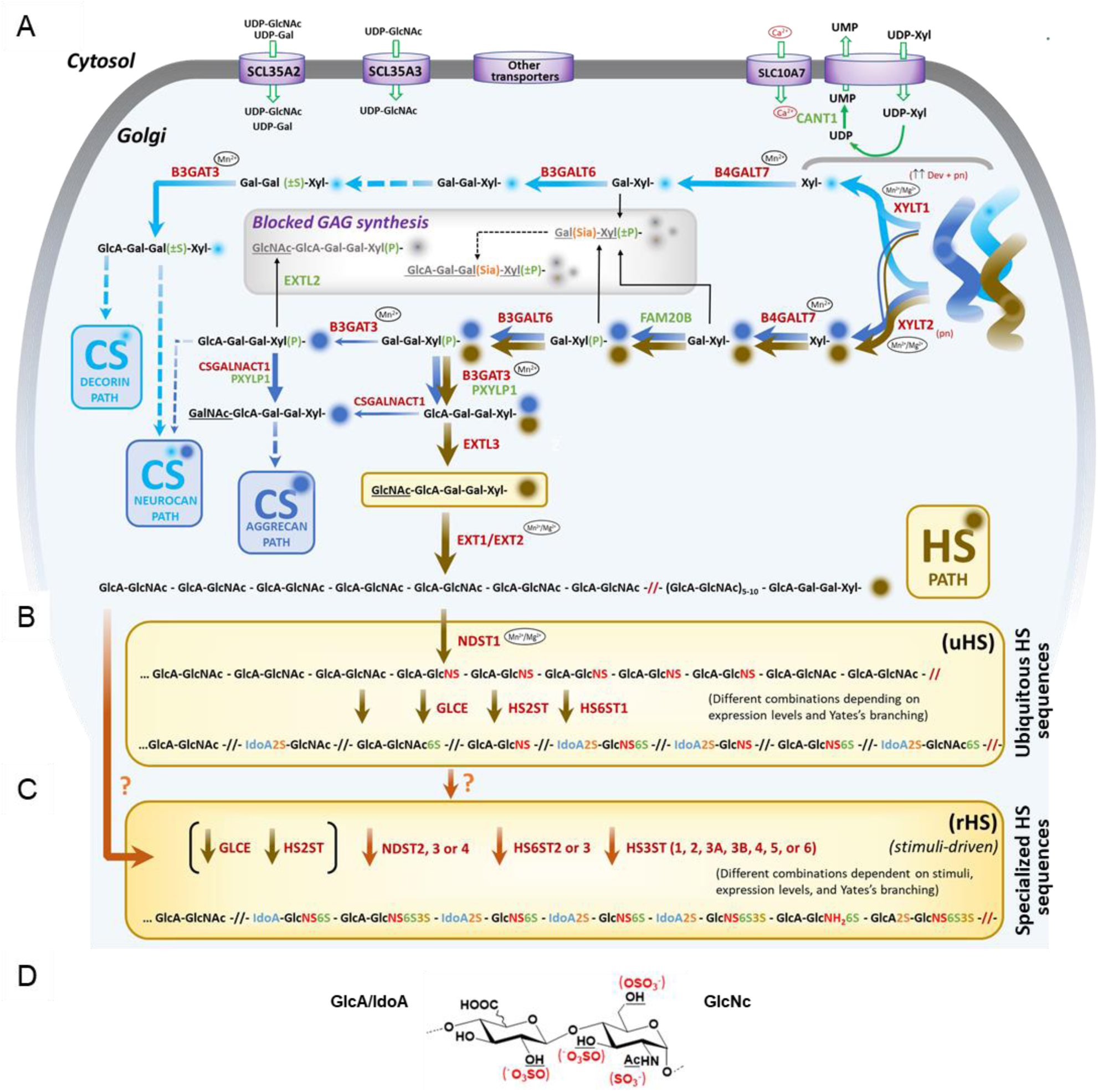
Schematic representation of the HSPG biosynthetic pathway. The HSPG path (brown arrows) overlaps with the CSPG-aggrecan path (marine blue arrows). The CSPG-decorin path (light blue arrows) is spliced from HSPG/CSPG path at early steps. Black arrows designate paths that stop GAG elongation. Arrow width designs preferred paths based in substrate preferences (see Gene review), wider arrows indicating preferred paths. Substrate pools guarantees maintenance of minor paths (narrow arrows). Dotted arrows indicate non-detailed steps. Green empty arrows indicate UDP-sugars or ion transport. Some nucleotide transporters or ion transports are represented. Ions as Ca^2+^, Mn^2+^ and/or Mg^2+^ are indicated when required for enzymatic activity. **A)** Biosynthesis of the GAG-CP linker tetrasaccharide common to HSPG and CSPG.), Excepting XYLT1 and XYLT2, all enzymes and transports are ubiquitously expressed during development (dev) and after birth (aft. birth). XYLT1 is highly expressed during development (**↑↑**dev) and its expression decreases aft. birth and preferentially binds to low molecular weight CSPG core proteins, as decorin (leading to favored CSPG-decorin path). XYLT2 is expressed aft. birth and shows similar binding to HSPG and CSPG core proteins (leading to both CSPG-decorin and HSPG/CSPG-aggrecan paths). Xyl is not phosphorylated in the CSPG-decorin path. **B)** Ubiquitously expressed enzymes drive the synthesis of ubiquitous HS sequences (uHS). **C)** Tissue restricted enzymes form HS sequences (rHS) likely during response to stimuli. rHS might contain high NS domains formed either from uHS or directly from unsulfated chains). **D)** Representative HS disaccharide indicating GlcA, IdoA and GlcN units and sulfation positions (in red).

**Table 3.** Ubiquitous heparan sulfates (uHS) biosynthetic genes. Genes are organized based on their expression in human tissues and on clinical variants traits and/or phenotype of the corresponding null mice.

|  | Gene | Impact of genetic variants in human | Phenotype of the null mouse | Most affected organs or tissues | Ref. |
| --- | --- | --- | --- | --- | --- |
| GROUP I<br>Essential<br>Widely expressed | *EXTL3* | Severe autosomal recessive skeletal dysplasia, including epispondylo-metaphyseal dysplasia, with developmental delay, immunodeficiency, and neuromotor and brain development delay. | Embryonically lethal at around 9 days post-fecundation with HS non-detected and with over production of CS in chondrogenic tissues. | All tissues. | 54,167 |
|  | *EXT1* | Hereditary multiple exostoses (multiple cartilaginous tumors), scoliosis, seizures, macrocephaly, defects in pancreas, lung, heart, cornea, etc. | *Ext1*<sup>-/-</sup>*Ext2*<sup>-/-</sup> mice die at embryo due to gastrulation failure. *Ext1*<sup>-/-</sup> form several exostoses. | Cartilage and bone. | 55,158 |
|  | *EXT2* | Hereditary multiple exostoses (less severe than that caused by EXT1). | *Ext1*<sup>-/-</sup>*Ext2*<sup>-/-</sup> mouse dies at embryo due to gastrulation failure. *Ext1*<sup>-/-</sup> mouse forms several exostoses. | Cartilage and bone. | 55,56, 158 |
|  | *NDST1* | Compound heterozygous mutations cause developmental delays, muscular hypotonia, ataxia, history of seizures, intellectual disability, epilepsy, gastroesophageal reflux, minor malformation, etc. Partially compensated by NDST2 except in brain (expressing NDST3 and 4), and skeletal muscle. | *Ndst1*<sup>-/-</sup> perinatally dies by respiratory failure and shows skeletal, brain, and heart defects. Heterozygous mouse shows severe developmental defects of the forebrain and forebrain-derived structures. | Brain, lung, skeletal and hearth muscles, stomach, etc. | 58,61 |
|  | *GLCE* | No pathogenic variants reported possibly because lethal. SNP associations with hypertension, and cerebrovascular events. | Neonatally lethal with severe developmental abnormalities on kidney, lung, skeleton, spleen, thymus, lymph node, etc. | Several organs: kidney, skeleton, thymus, lung, lymph node, spleen, etc. | 11,57, 168 |
|  | *HS2ST1* | No pathogenic variants reported possibly because lethal. SNP has been associated to n birds and pig has been associated with low metabolic rate and longevity in pigs and birds. | Neonatally lethal with onset of abnormalities after mid-gestation leading to traits including complete failure of kidney development. | Several tissues, particularly kidney. | 60,169 |
|  | *HS6ST1* | No pathogenic variants reported possibly because lethal. SNP association with delayed puberty and idiopathic hypogonadotrophic hypogonadism. | Lethal at late embryonic stages, with abnormalities in lung morphology, angiogenesis, and retinal axon guidance. If viable, mice showed growth retardation and lung defects. | Several organs: lung, kidney, cartilage, vascular system, reproduction organs, etc. | 59 |

#### 3.4.2. Other enzymes and transporters

In addition to variability in GAG-CP linker biosynthetic genes, genetic variability in ion and nucleotide transporters, or in other Golgian proteins that allow availability of sugar nucleotides and ions, can dramatically alter the biosynthesis of the GAG-CP linker^154,155^. For instance, the calcium activated nucleotidase-1 (CANT1) hydrolyses the uridine diphosphate nucleoside (UDP), which inhibits glycosyltransferases (as XYLTs)^155^ (Fig. 6A). Accordingly, *CANT1* clinical variants lead to Desbuquois dysplasia traits like those observed in *XYLT1* and *B4GAT7* variants^8,151^ (Table 2). Because CANT1 is calcium dependent, variants in the Ca^2+^ transporter SLC10A7 result in skeletal dysplasia with *CANT1* and *XYLT1* overlapping traits^10^ (Table 2). Similarly, variants in genes coding for sulfate ion transporters (*SLC26A2*)^156^, nucleotide (UDP-GlcNAc and UDP-Gal) transporters (*SCL35A2* and *SCL35A3*), or ions (Mg^2+^ and Mn^2+^) transporters required for proper enzymatic activities at the Golgi (Fig. 6A), can result in linkeropathies-like phenotypes^9,155,157^.

### 3.5. Essential and non-essential HS sequences

#### 3.5.1. Ubiquitous HS are essential for normal development and homeostasis

After biosynthesis of the GAG-CP linker, formation of the HS chain starts by the action of the glycosyltransferase EXTL3^2^, which adds a GlcNAc residue to the dephosphorylated GlcA-Gal-Gal-Xyl-(Ser-CP) product of B3GAT3/PXYLP1 (Fig. 6B). EXTL3 deficiency results in reduced HSPG and increased CSPG levels, confirming that abundant GlcA-Gal-Gal-Xyl-(Ser-CP) and GlcA-Gal-Gal-Xyl(P)-(Ser-CP) substrate pools respectively favor CSGALNACT1 and CSGALNACT1/PXYLP1 activities, thus driving CS initiation (Fig. 6A). Accordingly, lack of *Extl3* in mice causes embryonic lethality possibly due to the lack of HS chain synthesis. In human, biallelic mutations in the *EXTL3* gene result in a neuro-immuno-skeletal dysplasia syndrome^54^, suggesting an imbalanced HSPG/CSPG-aggrecan paths with decreased HS synthesis and increased syntheses of CSPG aggrecan and neurocan (Fig. 6A and 6B, Table 3, and Supp. Inf.). Following continuation of the HS chain synthesis, the EXT1/EXT2 hetero-oligomeric complex elongates the HS chain by alternated addition GlcNAc and GlcA to the GlcNAc-GlcA-Gal-Gal-Xyl-(Ser-CP) substrate, which initiates HS chain elongation^2^ (Fig. 6B). In *Ext1/Ext2* depleted mice, lack of HS leads to gastrulation failure and embryonic death^55,56^, agreeing the essentialness of HS chains carried by proteoglycans. In human, *EXT1* or *EXT2* variants lead to hereditary multiple exostoses (Table 3), characterized by the formation of cartilage-capped bony outgrowths^158^, suggestive of the accumulation of EXT1/EXT2 GlcNAc-GlcA-Gal-Gal-Xyl-(Ser-CP) substrate, which is the EXTL3 product. Thus, this in turn results in increased GlcA-Gal-Gal-Xyl-(Ser-CP) pool, which nourishes the CSPG-aggrecan path (Fig. 6A). Thus, in the context of GlcA-Gal-Gal-Xyl-(Ser-CP) substrate pool excess, CSPG-aggrecan can be accumulated in tissues in which its synthesis is rich, explaining the epiphyseal growth plates resulting in cartilage-capped bony outgrowths. On the continuation of HS elongation, HS chain maturation is assured by the ubiquitous NDST1, HS2ST, HS6ST1, and C5-epimerase (*GLCE*), which respectively assure basal *N*-deacetyl-*N*-sulfation, 2-*O*-sulfation, 6-*O*-sulfation, and GlcA/IdoA epimerization^2^. Interestingly, NDST1 and HS6ST1 substrate specificities suggest that, in association with HS2ST and GLCE, these enzymes can form sequences carrying NA/NS domains^59,159-163^ (Supp. Inf.). Indeed, NDST1 can generate HS domains with lower level of *N*-sulfation than NDST2^164^ while HS6ST1 shows higher selectivity for non-2-*O*-sulfated residues (hexA-GlcNS), as confirmed in the *Hs6st1* null mouse^59^. According with their essentialness, uHS made of NA/NS domains might then be those that interact and regulate HBP required for normal development and homeostasis^17,165^. Accordingly, *Ndst1*^*-/-*^, *Hs2st1*^*-/-*^, and *Glce*^-/-^ mice perinatally die with severe developmental abnormalities^57,60,61^, and the *Hs6st1*^*-/-*^ mouse cannot thrive^59^ (Table 3).

Moreover, no pathogenic variants are known in human for *HS2ST1, HS6ST1*, or *GLCE*, possibly because lethal. However, individuals with compound heterozygous *NDST1* mutations can survive, although with brain and muscle overt phenotypes^58^. This is possibly due to NDST2 redundance, as *NDST2* is not expressed in brain and skeletal muscle (Fig. 4)^58,166^. Together, these data support that uHS sequences synthetized by the Group I HSPG biosynthetic machinery could be those which, depending on their level of expression in each tissue, form HS sequences able to interact and regulate the activities of heparin binding proteins that assure tissue basal functions and homeostasis, as growth factors, morphogens, and other pericellular and cell membrane-associated proteins centrally involved in normal development and homeostasis.

#### 3.5.2. Non-essential HS in the appropriate response to stimuli

NDST2, NDST3, NDST4, HS6ST2, HS6ST3, and all HS3STs, are tissue restricted sulfotransferases coded by genes clustered in Group IV (Fig. 4, Fig. 5, and Suppl. Inf). No pathogenic variants are known for any of these genes and the corresponding knockout mice, when available, develops normally, are fertile, and live healthy (Table 4, and Supp. Inf.). Although it cannot be excluded that absence of deleterious phenotypes can be due to compensation phenomena, polymorphisms in most of these genes have been associated with resistance or risk to altered conditions affecting the organs in which they are expressed (Tables 4,see Suppl. Inf.). For instance, in brain, SNP in *NDST3* has been associated with risk of schizophrenia and bipolar disorders^99,101^, SNP in *NDST4* has been associated with reading difficulty and language impairment^102^, SNP in *HS6ST2* has been associated with intellectual disability^105^, SNP in *HS6ST3* has been associated with dysregulation of satiety^101,106^, and SNP in *HS3ST1* has been associated with to Alzheimer’s disease^108,110^. Moreover, the involvement of some of these genes in the appropriate response to neurological stimuli is starting to be documented. For instance, the *Ndst3*^*-/-*^ mouse exhibited subtle impaired anxiety with not compensatory effects from other *Ndst*^100^ while HS3ST2 expression was increased in the rat pineal gland after adrenergic stimulation^170^, indicating a role of these enzymes in adaptative neurological functions.

**Table 4.** Tissue restricted heparan sulfate (rHS) biosynthetic genes. Genes are organized based on their expression in human tissues, traits of clinical variants, and/or phenotype of the corresponding null mice.

|  | Gene | Impact of genetic variants in human | Phenotype of the null mouse | Most affected organs or tissues | Ref. |
| --- | --- | --- | --- | --- | --- |
| GROUP IV<br>Non-essential<br>Partial restricted expression | *NDST2* | No pathogenic variants reported. SNP association with risk of coronary artery disease and chronic kidney disease. | Viable, healthy, and fertile. Mast cell shows decreased sulfation in HS. | Mast cells | 176,177 |
|  | *HS6ST2* | No pathogenic variants reported. SNP association with risk of X-linked intellectual disability and myopia. | Viable, healthy, and fertile. | Brain and cartilage. | 105 |
|  | *HS6ST3* | No pathogenic variants reported. SNP association with decreased risk to obesity and to increased diabetic retinopathy. | Not reported. | Brain, eye. | 107,178 |
|  | *HS3ST1* | No pathogenic variants reported. SNP association with risk of arteriosclerosis, coronary artery diseases, and Alzheimer's disease. | Viable, healthy, and fertile but show proinflammatory phenotype when submitted to pathologic stimuli. | Vascular system and brain. | 179,180 |
|  | *HS3ST2* | No pathogenic variants reported. Expression is increased in Alzheimer's disease. SNP association with risk of schizophrenia. | Viable, healthy, and fertile. Responds to adrenergic response to light stimuli. | Brain. | 12,170, 181 |
|  | *HS3ST3A1* | No pathogenic variants reported. SNP association with risk of HSV1 and *P. falciparum* infection. | Not reported. | Vascular system. | 173,182 |
|  | *HS3ST3B1* | No pathogenic variants reported. SNP association with risk of *P. falciparum* infection. | Not reported. But overexpression affects immunity (peripheral T lymphocytes and Jurkat T cells adaptive response to stimuli). | Immune system. | 173,182, 183 |
| GROUP IV<br>Non-essential<br>High restricted expression | *NDST3* | No pathogenic variants reported. SNP association with risk of mental disorders as schizophrenia and bipolar disorders. | Develop normally, are fertile, and show only subtle hematological and behavioral abnormalities, with reduced anxiety. | Brain. | 99-101 |
|  | *NDST4* | No pathogenic variants reported. SNP association with risk of reading disability and language impairment. | No pathological outcomes, but some anomalies can be observed in histology images of the proximal colon, where the gene is not expressed . | In cartilage, brain, immune system. | 102,103 |
|  | *HS3ST4* | No pathogenic variants reported. SNP association with risk of altered verbal declarative memory, age-related macular degeneration, schizophrenia, and Alzheimer's disease. | Not reported. | Brain. | 116,117, 178,184 |
|  | *HS3ST5* | No pathogenic variants reported. SNP association with risk of intellectual disability and altered gray matter volume in schizophrenia. | Not reported. | Brain. | 117,185 |
|  | *HS3ST6* | No pathogenic variants reported. SNP association with risk of altered response to stimuli that triggers angioedema. | Not reported. | Skin, vascular system. | 118,119 |

In peripheral tissues, several polymorphisms have been associated, along or in combination with other genes, with altered conditions or susceptibility to disease (Table 4). For instance in coronary artery diseases (*HS3ST1*), ulcerative and Crohn’s colitis (*HS3ST2*)^111^, chronic obstructive pulmonary disease (*HS3ST2*)^112^, and hereditary angioedema (*HS3ST6*)^119^. The importance of these genes in the regulation of response to stimuli is also supported by available knockout mice and biomedical research. For instance, the *Hs3st1* null mouse showed a strong proinflammatory phenotype^109^, *HS3ST3B1* was shown to be involved in T lymphocytes activation^171^, *HS3ST3A1* expression was found increased during normotensive and pre-eclamptic pregnancies^113^, and *NDST2* was essential for mast cells responsiveness during inflammation and during innate and adaptive immunity toward pathogens^172^. Moreover, 3-*O*-sulfated HS (3S-HS) are well known to increase susceptibility to infection by pathogens^12^. Accordingly, polymorphism in both *HS3ST3A1 and HS3STB1* has been associated with risk to *P. falciparum* infection^173^ and some 3S-HS structures trigger HSV infection^12^.

#### 3.5.3. HS chain remodeling and biosynthesis arrest

After biosynthesis, HSPG are secreted to the cell surface or to the pericellular space, where their HS chains can be remodeled by SULF1, SULF2^14^, and/or by heparanase^15^, which activity can be negatively regulated by the enzymatically inactive heparanase 2^67^ (Table 5). Although no clinical variants are known for *SULF2*, the *Sulf2*^-/-^ mouse shows lethality^14^ (included in Group I), whereas in human, *HPSE2* variants result in pos-developmental syndromes (included in Group II), and other altered conditions (Suppl. Inf.)^68,174,175^. This indicates that both SULF2 and HPSE2 are required for normal homeostasis and basal response to stimuli. The involvement of HPSE2 in response to stimuli was confirmed in the homozygous *Hpse2* gene trap mouse, which developed normally but showed deficient responses to chemical and electrical field stimulation and showed low weight gain after weaning and died early in life^174,175^. In the other side, both *Sulf1*^-/-^ and *hpse*^-/-^ mice develop and live normally, are anatomically normal fertile, and show non overt phenotypes^14,186^ (included in Group III). Accordingly, non-pathogenic variants are known in human(Table 5). However, SNP has been associated with chronic graft-versus-host disease^84^, supporting its clustering in Group III. Accordingly, as for other genes in Group III, nonpathogenic *SULF1* variants are known, but polymorphism has been associated to multiple sclerosis^70^, fetus failure during *in vitro* fertilisation^88^, and preeclampsia^89^ (Table 5).

**Table 5.**
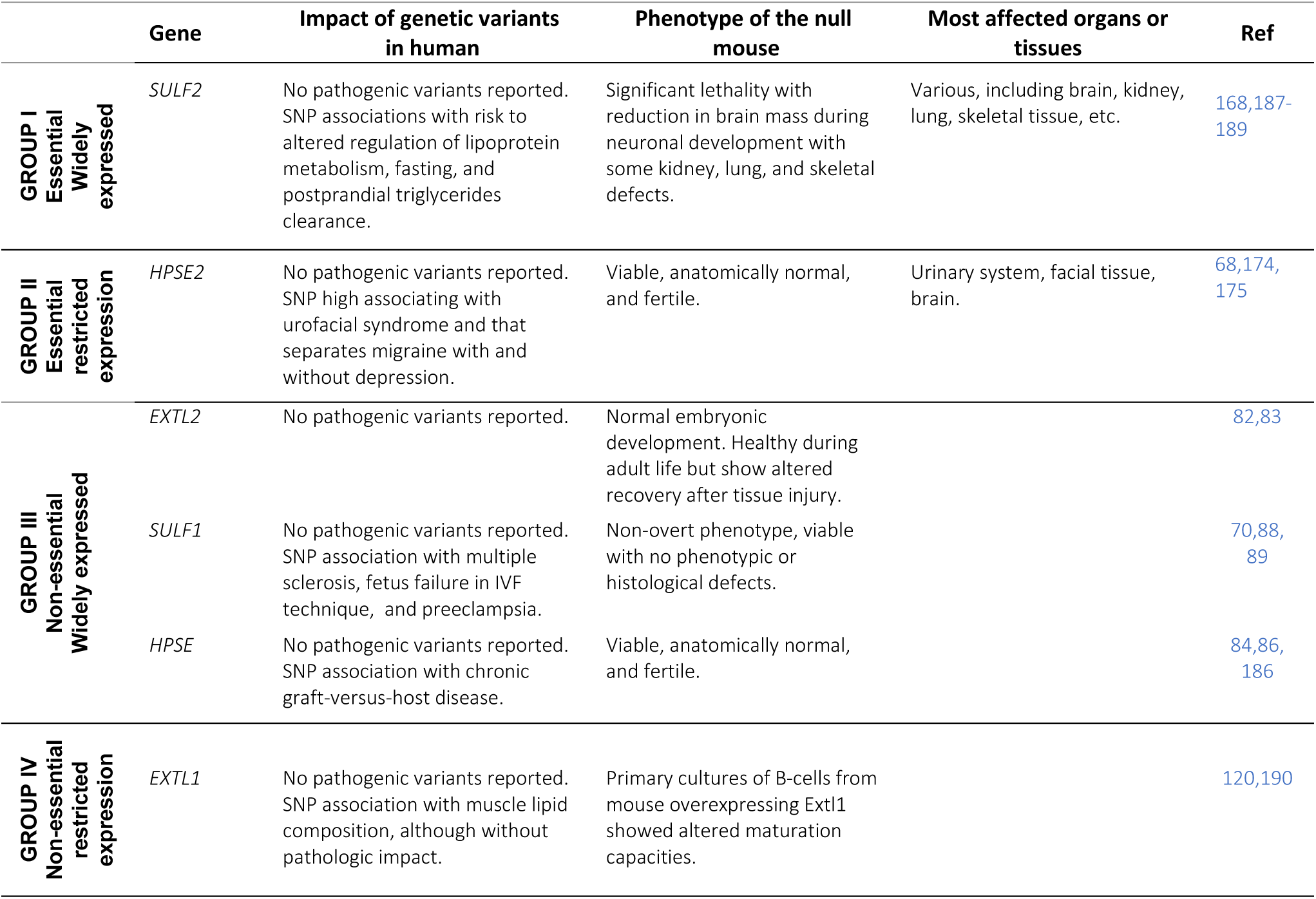
Genes coding for heparan sulfates remodeling enzymes. Genes are organized based on their expression in human tissues, clinical variants traits, and/or phenotype of the corresponding null mice.

Moreover, although widely distributed, *HPSE* and SULF1 basal expressions are low (Fig. 4), possibly because it is mainly required during tissue remodeling, as during wound healing, inflammation, and tumor growth and metastasis^15^. Interestingly, this is also the case for HPSE2 (Fig. 4), which main action could also be regulatory during endogenous stimuli. Together, these reports suggest that arrest of HS biosynthesis and HS chain remodeling predominantly occurs when cells and tissues are submitted to instigating or adaptative conditions, and that their SNP might predispose disease.

#### 3.5.4. Stimuli driven arrest of HS chain initiation or elongation

During biosynthesis, EXTL1 (Group IV) and EXTL2 (Group III) can control HS levels by respectively blocking chain elongation or chain initiation when the biosynthetic machinery is overactivated, for instance during wound healing, during adaptative immunological response of cells, or following other internal or external stimuli^13^. No pathogenic variants have been reported for neither of these genes and no genetic polymorphism has been associated with disease (Table 5). Indeed, to date, only polymorphism in *EXTL1* has been associated with muscle lipid composition, although without pathologic impact^120^. However, *in vitro*, primary cultures of B-cells from mouse overexpressing *Extl1* showed altered maturation capacities^190^ (Table 5), suggesting that but the enzyme might be involved in generation of environments or processed assuring an appropriate responsiveness to stimuli, including immunological stimuli^190^. Similarly, the *Extl2*^*-/-*^ mouse showed non overt phenotype, although presented impaired recovery following liver and kidney injury^82,83^. These data suggest that arrest of HS chain initiation or elongation is not essential during development and homeostasis but is required for appropriate response to stimuli and that SNP can predispose to altered conditions with or without clinical significance.

## 4. Discussion

HSPGs are among the more complex molecules in biology due to their HS structural and functional diversity. This diversity results from the complexity of their biosynthetic machinery coded by dozens of genes, which unclear relation to each other has hidden advances in the understanding of proteoglycans biology. Here, by following the PRISMA guidelines for systematic reviews and meta-analysis^18^, we retrieved information for the 50 main human HSPG biosynthetic genes, including those shared with the CSPG biosynthesis through the GAG-CP-linker tetrasaccharide, and for other genes involved in Golgian homeostasis. Analysis of the documented relationship between human pathogenic variants, SNPs associations with altered conditions or susceptibility to disease, null mice phenotypes, gene expression profiles, and substrates specificities of enzymes coded by the biosynthetic genes, resulted in the clustering in 4 Groups of genes organized depending on their essentialness and expression (Fig. 5). Globally, Group I integrates ubiquitous genes which by themselves can produce complete HSPG, as the group included genes coding for cores protein, all the GAG-CP linker biosynthetic enzymes, and glycosyltransferases and three sulfotransferases involved in the synthesis of a complete HS chain. Concerning the core proteins, Group I included genes coding for all the full time pericellular HSPG core proteins (perlecan, agrin, and collagen 18)^1^ and for 3 membrane-associated glypicans (glypicans 3, 4, and 6)^125^, indicating that this group can afford both essential pericellular HSPG involved structure and function of the pericellular matrix and and cell membrane-associated HSPG involved in cell signaling.

Concerning the GAG-CP linker tetrasaccharide coding genes, analysis of their clinical variants, null mice phenotypes, and substrate selectivities of their coded enzymes, resulted in a new organization of the GSG-CP linker biosynthetic pathway in which the HSPG synthesis is primarily assured over that of CSPG and in which the phenotypic continuum of disorders as linkeropathies and other pathologies is understandable^52,140,191^. Moreover, this new representation of the GAG-CP linker biosynthetic genes supports the idea that HSPG biosynthetic enzymes can assemble into ‘GAGosmes’^64,192^, or ‘Proteoglycanomes’^193^, which consider the integration of proteoglycan core proteins in the protein complexes that assure proteoglycans synthesis in the Golgi. Indeed, common protein modules in the ubiquitous/essential pericellular perlecan and agrin (Group I) are different from those shared by the ubiquitous/non-essential syndecans (Group III)^1^, but common to those in CSPGs aggrecan and neurocan, suggesting that agrin, perlecan, aggrecan, and neurocan can circulate through a common HSPG/CSPG path during GAG-CP linker synthesis, as proposed in Fig. 6A. Moreover, aggrecan and neurocan modules are different from those carried by decorin^1^, supporting the splitting of the CSPG-decorin path from the HSPG/CSPG-aggrecan path under interaction with XYLT1 at early steps of the GAG-CP linker biosynthesis (Fig. 6A). Likewise, syndecans modules are different from those carried by agrin, perlecan, aggrecan, neurocan, and decorin^1^, suggesting that the tissue restricted syndecans might integrate protein complexes different to those in which the ubiquitous core proteins are integrated. Thus, it will be important to investigate whether the tissue restricted HS biosynthetic enzymes in Groups III and IV contain protein modules that could associate them to syndecans or to other tissue restricted core proteins. Answering to this question will give information on whether rHS can be carried by uCP (Fig. 6C), as syndecans, but not only.

Concerning the HS chain, the EXTL3, EXT1, and EXT2 glycosyltransferases coding genes comprehensibly clustered in Group I, which contains genes that are essential for normal development and homeostasis (EXTL3, EXT1, and EXT2 are required for constructing the HSPG polysaccharide). Moreover, Group I clustered the ubiquitous *NDST1, GLCE, HS2ST1*, and *HS6ST1*, which appeared to be involved in the generation of uHS sequences that can then be those essential for normal development and homeostasis. In the other side, the tissue restricted *NDST2-4, HS6ST2-3*, and *HS3ST1-6* appeared to generate specialized rHS sequences mainly involved in responsiveness to stimuli. Based in their substrate specificities^59,159-163^, NDST1 and HS6ST1, in association with HS2ST and GLCE, likely form uHS NA/NS domains (Supp. Inf.), which sulfation level in each position might vary depending on these ubiquitous gene expression levels in the different tissues or cells^63-65^. Indeed, NDST1 generates HS domains with lower level of *N*-sulfation than NDST2^164^ while HS6ST1 shows higher selectivity for non-2-*O*-sulfated residues (hexA-GlcNS), as confirmed in the *Hs6st1* null mouse^59^. In the other side, mostly based in the presence of 3-*O*-sulfation and higher levels of *N*-and 6-*O*-sulfations that result from NDST2 *and* HS6ST2 substrate specificities^12,97,100,159,161-163,177,194-199^, it appeared that specialized rHS are formed by NS domains^12,162,177^. The existence of different synthetic paths leading to NA/NS or NS domains agrees the HS biosynthetic tree proposed by Yates *el al*. in which different enzymes specificities and substrate abundances can determine the HS synthesis direction either through a major branch assuring commonly occurring HS disaccharides or a minor branch assuring least common disaccharides^200^. Separation of the two branches seems to be associated to the relative abundance of two sets of structures resulting from the C5-epimerase preference to convert GlcA-GlcNS to IdoA-GlcNS compared to converting GlcA-GlcNAc to IdoA-GlcNAc^65,200^. Thus, because they are made by essential biosynthetic machinery, uHS NA/NS sequences might be those that are essential for normal development and homeostasis, whereas the specialized rHS sequences generated by enzymes coded by Group IV genes (*NDST2, NDST3, NDST4, HS6ST2, HS6ST3*, and all *HS3STs*) might be involved in specific biological functions. Accordingly, they are required for assuring adaptative behaviors and/or for proper responsiveness to instigating stimuli in the tissues or cells in which they are expressed.

### Ideas and Speculation

This work highlighted the possible existence of uHS likely tailored by NDST1, HS6ST1 in associating with HS2ST and GLCE, which together can form NA/NS domains. However, experimental evidence directly demonstrating that uHS sequences in ubiquitous HSPG are formed of NA/NS require to be established. Nevertheless, because of their essentialness, one can speculate that these uHS sequences are those that interact and regulate HBP required for normal development and homeostasis, including matrix proteins (collagen, laminin, etc.), growth factors (FGFs, VEGF, PDGF, BDNF, GDNF, etc.), morphogens (Wint, hedgehog, etc.), lipid binding proteins (apolipoproteins, lipoprotein lipase, etc.), cell adhesion proteins (P and L-selectin, integrins, etc.), among others^17,165^. Interestingly, several of these factors have been involved in cancer development and growth. Indeed, if NA/NS sequences are those that under physiological conditions interact with these factors and potentiated their activities, their excess might have over-signaling related oncogenic effects. Interestingly, SULF2 is a 6-*O*-sulfatase that shows strong pro-oncogenic activity and that also clustered in Group I. Thus, by de-sulfating NS sequences, SULF2 can generate biologically active NA/NS-like sequences that potentiate growth factors activities. If this is the case SULF2 will be pro-oncogenic, as is the case^14^. Although compensation phenomena among SULF1 and SULF2 can be argued, it noteworthy that while SULF2 is a pro-oncogene, SULF1 shows anti-oncogenic activity^14^, suggesting differential functions. Accordingly, SULF1 might inactivate excess of NA/NS sequences by de-sulfating uHS and thus generating under-sulfated 6-*O*-desulfated NA/NS domains unable to promote HBP trophic effects^14,62^. Thus, it is possible that by generating biologically active NA/NS-like sequences, SULF2 favors growth factors activities acting as pro-oncogene, whereas SULF1 participate to inactivation of active NA/NS sequences by generating under sulfated and inactive NA/NS domains, acting as anti-oncogene. This hypothesis requires large research to be confirmed or discarded. Similarly, based in information from the knockout mice and GWAS data, it appeared that specialized rHS sequences are those required for appropriate responsiveness to stimuli and adaptive behaviors in tissues in which each gene is specifically expressed. An additional question of interest is whether these specialized rHS are those that afford to HS chains their fine structural specificity, which will then not be directly related to fundamental processes in life, but mostly to specific tissue responsiveness to stimuli, resistance or vulnerability to disease or altered behaviors, as those observed in complex neurological disorders. Moreover, it is possible that rHS sequences (their biosynthetic genes knockout mice lives healthy) are carried by Group II and III core proteins (their biosynthetic genes knockout mice lives healthy), whereas uHS sequences (their biosynthetic genes knockout mice die) are carried by the ubiquitous core proteins (their biosynthetic genes knockout mice die). Answering to these questions will facilitate research aiming to understand the differential role of HSPG core proteins and the HS chains that they carry in a large variety of biological and pathological context. Thus, although to date it is not yet possible to sequence single HS chains isolated from their physiological contexts, the selective purification of core proteins from tissues or cells will make available their specific HS chains, in which the research of NA/NS or NS sequences could be performed. Finally, as HS chains are long, one should consider that the specialized rHS sequences might be those that gives to cells the capacity to generate specific HS interactomes with cell membrane proteins, or matrix proteins. The involvement of Group IV rHS biosynthetic genes in complex pathologies, as neurologic or behavioral disorders, might represent a solution for the identification of pathologies associating specific rHS interactomes. We are currently working in this direction.

**In conclusion**, by analyzing clinical variants and transcriptomic databases and literature records for biochemical and functional information on HSPG biosynthetic machinery genes, we decrypted essential features of proteoglycans and HS structural and functional diversity. Our analysis considers new concepts non developed in previous works, for instance those in which HS structures are defined by the HSPG biosynthetic genes expression levels^63-65^. From this analysis, we propose a clinical-variant and substrate selectivity-based proteoglycan biosynthetic pathway in which HSPG are primarily assured over CSPG, and in which the HS chains or sequences are characterized for carrying either an ubiquitous ‘sulfation level specificity’ (uHS) or a specialized ‘sulfation pattern specificity’ (rHS), depending on the tissue in where they are made. This work opens new routes of research for a better understanding of the roles of HSPG in physiology and pathology.

## Supporting information

Review on HSPG biosynthetic genes

HSPG biosynthetic genes

## Data Availability

All data produced in the present work are contained in the manuscript

## 5. Acknowledgments

This work has received funding from the ANR SkelGAG and from the European Union’s Horizon 2020 Research and Innovation Program (grant agreement No 737390). G. Barreto was funded by the “Centre National de la Recherche Scientifique” (CNRS, France), “Délégation Centre-Est” (CNRS-DR6), the “Lorraine Université” (LU, France) through the initiative “Lorraine Université d’Excellence” and the dispositive “Future Leader” and the “Deutsche Forschungsgemeinschaft” (DFG, Bonn, Germany) (BA 4036/4-1). We thank ArrestAD partners, particularly Prof. T. van Kuppevelt (Radboud University, the Netherlands) and Prof. D. Fernig (University of Liverpool, UK), to Prof. P. Albanese, Dr. A. Fifre (University Paris Est Créteil, France), Prof. J-P. Li, Prof. U. Lindahl, and Prof. L. Kjellen (Uppsala University, Sweden), for interesting discussions.

## 6. Competing interests

The authors declare that they have no competing interests.

## 7. Author contributions

DPG designed the study and wrote the manuscript. DPG, MOO, BDSF, and JDR analyzed the data. DPG, GB, BDSF, and MMO participated to data organization. DPG, MOO, GB, NR, and AM participated to figures design. SC, MBH, XL, GLD, and OGV participated to data analysis and literature review. All authors discussed results and commented the manuscript for improvement.

## Notes

### Competing Interest Statement

The authors have declared no competing interest.

### Author Declarations

For this study we used ONLY openly available human data that were originally located at : 1) the ClinVar archive (database ClinVar: https://www.ncbi.nlm.nih.gov/clinvar/intro/), which gives information on correlations between genetic variations and overt phenotypes or health status with history of interpretations, 2) the Orphanet archive (Database Orphanet: https://www.orpha.net/), which considers clinical presentation based on published scientific articles and expert reviews, and 3) the dbSNP archive (database: https://www.ncbi.nlm.nih.gov/snp/), which is a free public archive for genetic variation within and across different species. For clinical variants, only monogenic variants (indel, deletions, duplications, insertions, and single nucleotide) were considered. Clinical significance was further confirmed in web platforms including Online Mendelian Inheritance in Man (OMIM) (https://www.omim.org/) and the Database of Genomic Variants Archive (DGVa database: https://www.ebi.ac.uk/dgva/). For transcript analysis we used ONLY openly available human data that were originally located at : 1) the RNA-seq 32Uhlen project (database 32Uhlen: http://www.proteinatlas.org/humanproteome), which analyzed 32 different tissues from 122 human individuals, 2) the RNA-Seq CAGE (Cap Analysis of Gene Expression) in the RIKEN FANTOM5 project (database FANTOM5: http://fantom.gsc.riken.jp/data/), which analyzed several healthy adult human tissues, and 3) the ENCODE strand-specific RNA-seq of 13 human tissues from Michael Snyder's lab (Database ENCODE: https://www.encodeproject.org/). All these RNA-seq databases are considered in the expression atlas (http://www.ebi.ac.uk/gxa). The original raw and processed data files can be found in the ArrayExpress platform (https://www.ebi.ac.uk/arrayexpress/).

### Summary of Updates

Some errors corrected including in title, Figure 6 number, and other minor typo.

