## Supplementary material for "Variability in proteoglycan biosynthetic genes reveals new facets of heparan sulfates diversity. A systematic review and analysis": Review on HSPG biosynthetic genes

### HSPG biosynthetic genes review

The HSPG biosynthetic machinery genes were here reviewed following their allocation to Groups I, II, III, or IV. PRISMA guidelines[1] were used to search information on clinical variants for each human gene and for the corresponding null mouse. A second campaign was performed to retrieve information on each gene product functionally, including enzymes substrate selectivity.

Page

**GROUP I. Essential wildly expressed HSPG biosynthetic genes**

Group I – Essential widely expressed HSPG core proteins coding genes

*HSPG2* ..………………………………………………………….………… 3
*AGRN* ……………………………………………………….…………. 3
*COL18A1* ………………………………………………………………… 4
*GPC3* ……………………………………………………….……………... 4
*GPC4* ……………………………………………………….……………... 5
*GPC6* ……………………………………………………….……………... 5

Group I – Essential widely expressed GAG-CP linker biosynthetic genes

*XYLT1* ……………………………………………………….…………….. 6
*XYLT2* ……………………………………………………….…………….. 7
*B4GALT7* ……………………………………………………….………… 8
*FAM20B* ……………………………………………………….………... 8
*B3GALT6* ……………………………………………………….………… 9
*B3GAT3* ……………………………………………………….………... 10
*PXYLP1* ……………………………………………………….………... 11
*CANT1 and SLC10A7* ……………………………………….………... 11
*SLC26A2, SCL35A2,* and *SLC35A3* …………………………………... 12

Group I – Essential widely expressed HS chain biosynthetic genes

*EXTL3* ……………………………………………………………………. 13
*EXT1* and *EXT2* ……………………………………………………………. 13
*NDST1* ……………………………………………………………………... 14
*GLCE* ……………………………………………………………………... 14
*HS2ST1* ……………………………………………………………………... 15
*HS6ST1* ……………………………………………………………………... 16

Group I – Ubiquitous HS chain remodeling machinery genes:

*SULF2* ……………………………………………………………………... 16

**GROUP II. Essential tissue restricted HSPG biosynthetic genes**

Group II – Essential tissue restricted HS remodeling control:

*HPSE2* ……………………………………………………………………. 17

**GROUP III. Non-essential wildly expressed HSPG biosynthetic genes**

Group III - Non-essential widely expressed HSPG core proteins

*GPC1* ……………………………………………………………………... 18
*SDC1* ……………………………………………………………………... 19
*SDC2* ……………………………………………………………………... 19
*SDC3* ……………………………………………………………………... 20
*SDC4* ……………………………………………………………………... 20

Group III – Non-essential widely expressed HS chain remodeling genes

*EXTL2* ……………………………………………………………………... 20
*SULF1* ……………………………………………………………………... 21
*HPSE* ……………………………………………………………………... 21

**GROUP IV. Non-essential tissue restricted HSPG biosynthetic genes**

Group IV – Non-essential tissue restricted HSPG core proteins

*GPC2* ……………………………………………………………………... 22
*GPC5* ……………………………………………………………………... 22

Group IV – Non-essential tissue restricted specialized HS chain maturation

*NDST2* …………………………………………………………………… 23
*NDST3 and NDST4* ……………………………………………………... 23
*HS6ST2* ……………………………………………………………………. 24
*HS6ST3* ……………………………………………………………………..... 24
*HS3ST1* ……………………………………………………………………. 25
*HS3ST2* ……………………………………………………………………... 26
*HS3ST3A1* ……………………………………………………………………. 26
*HS3ST3B1* ……………………………………………………………………. 27
*HS3ST4* ……………………………………………………………………... 27
*HS3ST5* ……………………………………………………………………. 28
*HS3ST6* ……………………………………………………………………. 28

Group IV – Tissue restricted HS chain remodeling genes

*EXTL1* ……………………………………………………………………. 29

#### GROUP I. Essential wildly expressed HSPG biosynthetic genes

##### Group I – Essential wildly expressed HSPG core proteins coding genes

*HSPG2* (Chr.1; ID:3339)

Heparan sulfate proteoglycan 2 (*HSPG2*) is a widely expressed (Fig. 4) and highly conserved gene that codes for the secreted perlecan core protein, which can carry over four long chains of HS[2]. Perlecan is the major HSPG in most basement membranes, which are the highly specialized extracellular matrices that tie the epithelium to connective, endothelial, muscular, neuronal, and other tissues[3]. During the early stages of embryogenesis, perlecan deposits in vascularized tissues such cartilage primordia, pericardium, blood vessels walls, and heart[2]. At later stages of development, perlecan is widely expressed in other tissues including lung, kidney, liver, gastrointestinal tract, and brain[4]. The highest deposition of perlecan occurs in tissues that are rich in extracellular matrix, as in cartilage undergoing endochondral ossification[4]. In human, *HSPG2* pathogenic variability leads to bone dysplasia and to several other defects including cardiovascular and pulmonary phenotypes, myotonia, and hyperthermia. These defects range from relatively mild conditions (Stuve-Wiedemann syndrome and the Schwartz–Jampel syndrome) to severe and/or lethal diseases (Lethal Kniest-Like Syndrome and Dyssegmental Dysplasia of Silverman-Handmaker type)[2], with a degree of severity inversely correlating with the amount of perlecan being deposited into the extracellular matrix[5]. Accordingly with the most severe phenotypes, *Hspg2*$^{-/-}$ mice perinatally die with skeletal dysplasia and/or defective cephalic development and several other traits[4]. The perlecan phenotype in the knockout mice agrees its biological role in the pericellular space in where the proteoglycan binds, cross-links, and stabilizes many molecules, including extracellular matrix structural proteins, growth factors, morphogens, and cell-surface receptors[5]. Because of its essential role in normal embryonic development and its wild tissue distribution, *HSPG2* was HERE allocated to the Group I of HSPG biosynthetic machinery genes (Fig. 4 and Fig. 5).

*AGRN* (Chr.1; ID:375790)

*Agrin (AGRN)* is a widely expressed gene (Fig. 4) that codes for the secreted core protein agrin, predominantly present in the basement membranes that join muscle fibers and nerves at the neuromuscular junction[3], although the protein is also present in other basement membranes and extracellular matrices[3,6]. In agreement with its essential role in normal embryonic development, particularly at the neuromuscular junction, *AGRN* clinical variants cause congenital myasthenic syndromes including distal muscle weakness and atrophy[7], and SNP has been associated to myasthenia gravis[8], a rare autoimmune disorder affecting the neuromuscular junction. Accordingly, *Agrn*$^{-/-}$ mice died at birth due to respiratory failure and central nervous system defects[6,9]. The involvement of agrin in the organization of basement membranes other than those in the neuromuscular junction was revealed by the selective re-expression of the gene in motor neurons of the *Agrn*$^{-/-}$ mice, in which *Agrn* re-expression could prevent perinatal death, although mice exhibited severe postnatal growth retardation with markedly altered growth plate morphology, consistent with the implication of this HSPG in basal membranes during musculoskeletal development[3,6]. Because of its essential role in embryonic development and wide distribution in basement membranes, particularly at the neuromuscular junction, *AGRN* was allocated to the Group I of HSPG biosynthetic machinery genes (Fig. 4 and Fig. 5).

*COL18A1* (Chr 21; ID:80781)

Collagen XVIII is a secreted HSPG core protein coded by the ubiquitously expressed *COL18A1* gene, which is predominantly expressed in basement membranes of epithelial and vascular tissues [10] (Fig. 4). In human, 3 collagen XVIII variants differing in their *N*-terminal non-collagenous sequences are known, all of them carry at least three HS chains. Mutations in *COL18A1* result in Knobloch syndrome, a genetic disorder characterized by severe eye defects (e.g. myopia, vitreoretinal degeneration, macular abnormalities) and occipital encephalocele[11]. Accordingly, lack of the collagen XVIII in the *Col18a1*$^{-/-}$ mice results in a deleterious phenotype affecting eye and encephalocele[12], although it also affects base membranes of many other tissues including kidney proximal tubules, heart valves, epidermis, choroid plexus[13], as supported by increasing evidences in situations such as atherosclerosis, glomerulonephritis, and in other type of tissue damage[10,11] in which collagen XVIII seams to participate to the regulation of cell survival, stem or progenitor cell maintenance and differentiation during inflammation. Because of its extensive tissue distribution and of the sever overt phenotypes observed in human clinical variants and knockout mice, *COL18A1* was allocated to the Group I of HSPG biosynthetic machinery genes (Fig. 4 and Fig. 5).

*GPC3* (Chr.X; ID:2719)

*GPC3* encodes the widely expressed (Fig. 4) membrane-associated glypican 3 core protein[14], which clinical variants are responsible of the X-linked Simpson-Golabi-Behmel syndrome, characterized by pre and post-natal overgrowth with skeletal, facial, visceral, and neurological anomalies[15]. These abnormalities are recapitulated in the *Gpc3*$^{-/-}$ mice, which exhibited perinatal death with developmental dysmorphisms including abnormal lung and kidney[16]. Other *GPC3* variants are responsible of Wilms tumor 1 (nephroblastoma), the most common kidney cancer in children, and with hepatocellular carcinoma[17]. Because of its high levels, glypican 3 has been proposed as biomarker and target for nephroblastoma and hepatocellular carcinoma[18]. Indeed, glypican 3 plays important biological roles by interacting and regulating the function of HBP including BMPs, WTNs, SHHs, and FGFs, among others[17]. As other glypicans, glypican 3 carries a *N*-terminus signal domain, a globular intermediary domain containing multiple cysteine residues, and a C-terminus domain carrying the GPI that anchors the core protein to the cell surface. HS chains are attached to a C-terminus short domain that keeps the sugar chain close to cell membrane areas rich in lipids, allowing the concentration of HBP near to their high affinity cell surface receptors that allow cell signaling. Because of the overt phenotype of its genetic variants in human and knockout mice, *GPC3* was allocated to the Group I of HSPG biosynthetic machinery genes (Fig. 4 and Fig. 5).

*GPC4* (Ch.X; ID:2239)

*GPC4* encodes the membrane associated HSPG glypican 4 core protein, widely expressed during development and adulthood (Fig. 4). Interestingly, *Gpc4* was found enriched in the hippocampus of the developing mice, suggesting a central involvement in brain development[19]. In human, *GPC4* pathogenic variants cause Keipert Syndrome, characterized by prominent forehead, flat midface, hypertelorism, broad nose, digital abnormalities, and deafness, indicating a role of this HSPG in the regulation of morphogen activities and in perceptive impairment[20]. Accordingly, the *Gpc4*$^{-/-}$ mice reproduce the main traits of Keipert syndrome, including craniofacial and digital abnormalities[20], as well as behavioral alterations associated to abnormal levels of GluA1 and AMPARs at the synapsis[21]. In agreement with its implication in cognition and social behavior, an altered expression of *GPC4* has been linked to autism spectrum disorders (ASD)[21,22]. Because of its wide distribution in tissues and the deleterious morphogenic and cognitive phenotypes in human clinical variants and knockout mice, *GPC4* was allocated to the Group I of HSPG biosynthetic machinery genes (Fig. 4 and Fig. 5).

*GPC6* (Ch13; ID:10082)

*GPC6* is a widely expressed gene that encodes the HSPG glypican 6 core protein[19] (Fig. 4). In human, *GPC6* loss-of-function mutations result in an autosomal-recessive condition known as omodysplasia 1 (OMOD1), characterized by facial dysmorphism, short stature, and shortened limbs[23]. In agreement with its requirement for normal embryonic development, *Gpc6*$^{-/-}$ mice die at birth with severe facial dysmorphism and bones significantly shorter than those of their normal littermates. This phenotype has been related to a signaling deficit of the morphogenic HBP sonic hedgehog[24]. Interestingly, a role of *GPC6* during adult bone homeostasis is currently supported by genetic wide association studies (GWAS) showing that SNP adjacent to the *GPC6* coding sequences is a risk factor for osteoporosis[25]. Moreover, altered *GPC6* expression has been implicated in melanoma[26] and prostate cancer[27], indicating a role of glypican 6 in tumor progression possibly through its capacity to assure growth factors cell signaling[17]. Because of the deleterious phenotypes of its clinical variants in human and lethality of the knockout mice, *GPC6* was allocated to the Group I of genes involved in HSPG biosynthesis (Fig. 4 and Fig. 5).

**Group I – Essential wildly expressed GAG-CP linker biosynthetic genes**

Based on their wide expression profiles and overt phenotypes caused by clinical variants in human and in knockout mice (Fig. 4), the GAG-CP linker tetrasaccharide biosynthetic genes *XYLT1*, *XYLT2*, *B4GALT7*, *FAM20B*, *B3GALT6*, *B3GAT3*, *PXYLP1*, *CANT1* and *SLC10A7*, were clustered in Group I. Because the GAG-CP linker tetrasaccharide supports both HS and CS chains in proteoglycans, it would be expected that defects in the GAG-CP linker biosynthetic machinery will lead to phenotypes associated to both GAGs, unless a preferred pathway favor the synthesis of a proteoglycan type over the other. To evaluate this possibility, we paid particular attention to literature reports on substrates specificities and products of the enzymes involved in the GAG-CP linker biosynthetic machinery. This analysis indicates that different alternative paths coexist during the GAG-CP biosynthesis, and that although some of these paths overlap, the biosynthesis of HSPG is preferably assured over that of CSPG. This can explain why the primarily observed phenotypes affect tissues classically rich in the non-preferred CSPG products. Interestingly, synthesis of the linker tetrasaccharide that will carry CS chains is itself spliced in different paths (Fig. 6A), likely assuring the biosynthesis of different CSPG types (not deeply treated in this analysis). The predominance of some pathways over others was inferred from data on substrate specificities and from the clinical traits associated to genetic variability on each biosynthetic enzyme, as reviewed in the next paragraphs.

*XYLT1* (Chr.16; ID:64131)

Xylosyl transferase 1, coded by *XYLT1*, is a widely expressed ion binding enzyme (Fig. 4), which at the cis Golgi catalyzes the transfer of Xyl from the UDP-Xyl donor to specific serine residues of the core protein (Ser/CP)[28,29]. During development, *XYLT1* is highly expressed, particularly in chondrocytes, which are the skeletal precursor cells that produce high levels of CSPG within the cartilage template that forms bones[28-30]. The critical role of the highly expressed *XYLT1* during cartilage template ossification is suggested by the severe skeletal phenotype observed in humans carrying *XYLT1* mutations, which are responsible of the skeletal Desbuquois dysplasia type 2 (DBQD-2), also called Baratela–Scott syndrome. DBQD-2 is characterized by short stature, advanced carpal ossification, distinct facial features, joint laxity, and intellectual disabilities[30,31]. Analysis of GAGs from primary fibroblasts of DBQD-2 patients have shown an important reduction of large CSPG, as aggrecan, and unchanged HSPG levels, indicating maintenance of HSPG path over the secreted large CSPG-aggrecan (Fig. 6A). Reproduction of one of the patient’s mutation in a cell line confirmed the reduction of large CSPG without affecting neither HS levels, nor small CSPGs (as decorin) levels. Indeed, XYLT1 interacts with decorin with higher affinity than with aggrecan, suggesting that XYLT1 can more efficiently transfer Xyl to Ser residues in the small-CSPG core protein than in the large one[32,33]. Accordingly, independent kinetic studies have shown a XYLT1 higher affinity for small-CSPG core proteins, whereas XYLT2 has similar affinity for small and large cores in both CSPG and for HSPG[29,30]. This suggests that clinical variants resulting in reduced XYLT1 activity during development, when XYLT2 is not expressed, will mainly affect large-CSPG (as aggrecan), leading to a strong chondrocytic phenotype. Although the *Xylt1*$^{-/-}$ mouse has not been generated possibly because of failure to develop, a pug mouse in which a homozygous missense mutation in *Xylt1* was produced by a *N*-ethyl-*N*-nitrosourea mutagenesis screen, showed a strong skeletal phenotype with premature maturation of chondrocytes, early ossification, and disproportionated dwarfism[28]. Summarizing, XYLT1 can assure synthesis of both HSPG and CSPG, although with preferred production of small-CSPG and HSPG. As *XYLT1* is critically required for normal embryonic development and its genetic variability results in deleterious phenotypes in human and mice[31], this gene was placed in the Group I of GAG-CP linker biosynthetic machinery genes common to HSPG and CSPG (Fig. 4 and Fig. 5).

XYLT2 (Chr.17; ID:64132)

Xylosyl transferase 2 (XYLT2), coded by *XYLT2*, is an ion binding protein expressed at post developmental stages. XYLT2 participates to the production of the Xyl-(Ser/CP) pool after birth[29], when XYLT1 expression decreases. Whereas *XYLT1* mutations cause severe developmental defects, mutations in *XYLT2* only cause post-developmental disorders, as spondyloocular syndrome, an autosomal recessive condition characterized by osteoporosis, cataracts, sensorineural hearing loss, learning difficulties, and atrial septal defect that appears after birth[31,34], translating deficiency of small-CSPG (decorin and neurocan paths affected) (Fig. 6A). Accordingly, *Xylt2*$^{-/-}$ mice display no developmental defects, but show post-natal overt phenotypes particularly in organs in which *Xylt1* expression is the lower, as in liver and adipose tissues (Fig. 4)[32,33], but not only[35]. XYLT2 shows a similar preference for the small and large CSPG and HSPG core proteins[28-30] (Fig. 6A). Kinetically, XYLT2 glycosylates HSPG and CSPG core proteins with similar efficiency[30], suggesting that after birth XYLT2 equality assures the production of Xyl-(Ser/CP) pool, when *XYLT1* expression is reduced to basal levels (Fig. 6A). After birth, XYLT2 can then participate to HSPG synthesis in organs in which the XYLT1 expression is the lowest[32,35]. Interestingly, XYLT2 activity can be measured in serum of patients with pathologies as systemic sclerosis and for the diagnosis of fibrotic remodeling processes [31], suggesting a weak XYLT2 anchoring to the Golgi membrane, although the physiopathological implication of this event remains to be investigated. Currently, several single *XYLT2* pathogenic variants are reported, although none is lethal, suggesting that XYLT1 can assure CP-Ser xylosylation in most organs. The overt phenotype observed in mice lacking *Xylt2* and its pathogenic variants in human place *XYLT2* in the Group I of HSPG (and CSPG) biosynthetic machinery genes essential for normal embryonic development and physiology (Fig. 4 and Fig. 5).

*B4GALT7* (Chr. 5; GeneID:11285)

*B4GALT7* encodes the ubiquitous cis-Golgi manganese ion binding β-1,4-galactosyltransferase 7 (B4GALT7) (Fig. 4), also known as galactosyltransferase I (GALT-I), which shows high UDP-galactose (UDP-Gal) substrate specificity[36]. B4GALT7 transfers the first galactose (Gal-I) to the growing Xyl-(Ser/CP) to form Gal-Xyl-(Ser/CP) (Fig. 6A). Because B4GALT7 is the only enzyme that uses the Xyl-(Ser-CP) pool, synthetized by XYLT1 from early developmental stages, the *B4galt7* knockout mice cannot be generated and genetic variability in human can be expected to result in a *XYLT1*-like developmental phenotype, although with stronger effect in the small-CSPG compared to *XYLT1*-variants, particularly after birth, when XYLT1 decreases. Accordingly, in human, compound heterozygous mutations in *B4GALT7* cause B4GALT7-related Ehlers-Danlos syndromes (B4GALT7-EDS), previously known as spondylodysplastic EDS type 1 (spEDS-1), characterized by alterations of connective tissue with joint hypermobility, skin hyperextensibility, and tissue fragility. In severe phenotypes, B4GALT7-EDS additionally shows short stature (skeletal tissue affections), muscle hypotonia, and other conditions which severities depend on the clinical variant[37,38]. One of the more severe forms is Reunion Island Larsen-like syndrome (RILLS), characterized by sdEDS-1 traits with additional trials including multiple dislocations, facial dysmorphism, ocular abnormalities, motor and cognitive delays[37-39], suggesting developmental alterations in both small- (decorin and neurocan) and large-CSPG paths, affecting HSPG at lower extent. Indeed, the clinical significances of *B4GALT7* variants range from benign to pathogenic[37,38] depending on the different catalytic efficiencies of the mutated enzyme[36,40], with the more severe traits likely correlating with decreased HS levels, although this remains to be confirmed. Thus, because of its requirement for normal embryonic development, *B4GALT7* was placed in the Group I of HSPG (and CSPG) biosynthetic machinery genes (Fig. 4 and Fig. 5).

*FAM20B* (Chr.1; ID: 9917)

The golgian xylosylkinase FAM20B (*FAM20B*) is a member of the ubiquitously expressed ‘Family with sequence similarity 20’ (FAM20) proteins (Fig. 4). FAM20B adds a phosphate group on the C2 of Xyl in Gal-Xyl-(Ser/CP) to form Gal-Xyl(P)-(Ser/CP), which is the preferred, although not exclusive, B3GALT6 substrate that leads to Gal-Gal-Xyl(P)-Ser/CP, the preferred B3GAT3 substrate [41,42] (Fig. 4A). Phosphorylation of Xyl occurs in proteoglycans that follow the common HSPG/CSPG path, whereas the unphosphorylated Xyl is characteristic of the small-CSPG decorin path. Accordingly, the Xyl phosphorylated linker tetrasaccharide has only been observed in HSPGs and in some CSPGs including aggrecan and neurocan, but not in decorin[40,42] (Fig. 6A). Therefore, FAM20B has been proposed to act as a molecular switch that outline the identity of the GAG chain that will be elongated in the GAG-CP linker [42]. To date, there is only one reported *FAM20B* compound heterozygous variant, which affected two siblings causing the death of one of them shortly after birth, the second one died at early infancy[43]. The two siblings showed a severe phenotype like *XYLT1*-Desbuquois dysplasia without joint laxity and very short stature, multiple dislocations of the large joints, mid-face, and thoracic hypoplasia with respiratory failure[43]. This phenotype evokes a defective HSPG/large-CSPG path (as aggrecan and neurocan)[40], but not defective small-CSPG decorin path[40], as proposed in Fig. 6A. More sever phenotypes evoking disruption of HSPG in the HSPG/large-CSPG path are evoked by the embryonal lethality of *Fam20b*$^{-/-}$ mouse, which showed sever stunted growth, multisystem organ hypoplasia and a delayed development most evident in the skeletal system, eyes, lung, gastrointestinal tract, and liver. Therefore, *FAM20B* was here allocated to the Group I of HSPG (and CSPG) biosynthetic machinery genes essential for normal embryonic development and tissue homeostasis (Fig. 4 and Fig. 5).

*B3GALT6* (Chr.1; ID: 126792)

The β-1,3-galactosyltransferase 6 (B3GALT6), also known as galactosyltransferase II (GALTII), encoded by the *B3GALT6* intron-less gene, is an enzyme present in the medial Golgi, where it catalyzes the transfer of the second Gal (Gal II) to Gal-Xyl(±P)-(Ser/CP) to produce Gal-Gal-Xyl(±P)-(Ser/CP), with higher selectivity for the phosphorylated substrate[41,42], favoring the HSPG/large-CSPG path (Fig. 6A). As B3GALT6 can produce both the phosphorylated and non-phosphorylated GAG-CP linker intermediates, it can act in both the HSPG/CSPG path and the small-CSPG decorin path (Fig. 6A). Accordingly, cultured fibroblasts from patients carrying biallelic *B3GALT6* mutations showed reduced production of both HSPG and CSPG[44,45]. In human, *B3GALT6* mutations cause a wide phenotypic spectrum similar to that associated to *B4GALT7*, so called B3GALT6-related spondylodysplastic Ehlers-Danlos syndrome (B3GALT6-spEDS2 or spEDS-2) or spondyloepimetaphyseal dysplasia with joint laxity[46]. As other EDS, these conditions affect connective tissues and skeletal tissues, including bone, skin, cartilage, tendons, and ligaments. Indeed, depending on the genetic variant, B3GALT6-spEDS2 shows overt phenotypes with variable severities involving joint hypermobility, short stature, muscle hypotonia, kyphoscoliosis, craniofacial defects, hyperextensible skin, delayed motor and/or cognitive development, joint contractures, ascending aortic aneurysm, and spondyloepimetaphyseal dysplasia[44,45]. Accordingly, analysis of patients with compound heterozygous mutations showed reduced levels of the B3GALT6 transcript or protein, while others show protein mislocalisation. Interestingly, deficient B3GALT6 activity was recently shown to produce a non-canonic linker tetrasaccharide lacking the second Gal moiety but able to elongate GAG chains, although with altered structures[47]. Although *B3galt6* knockout mice have not been produced, possibly because a defective HSPG/CSPG path incompatible with development and life, and because of the deleterious phenotype of *B3GALT6* genetic variants in human, this gene was placed in the Group I of HSPG (and CSPG) biosynthesis machinery genes (Fig. 4 and Fig. 5).

*B3GAT3* (Chr.11; ID: 26229)

*B3GAT3* codes for β-1,3-glucuronyltransferase 3 (B3GAT3), also known as glucuronosyltransferase I (GLCATI), a ubiquitous metal ion binding enzyme which at the cis Golgi catalyzes GlcA transfer from UDP-GlcA into Gal(±S)-Gal-Xyl(±P)-(Ser/CP) to form GlcA-Gal(±S)-Gal-Xyl(±P)-(Ser/CP) in both the HSPG/CSPG and the small-CSPG paths (Fig. 6A) . The critical role of this enzyme in development was confirmed by the early sever lethal phenotype observed during the *B3gat3*$^{-/-}$ mouse generation, which resulted in embryonic lethality before the 8-cell stage due to failed cytokinesis with strong reduction of the synthesis of both HS and CS chains[46,48]. In human, several *B3GAT3* clinical variants have been associated to *B3GAT3*-related syndromes affecting connective, skeletal, cardiac, and other tissues, with severities depending on the variant. As B3GAT3 is involved in the different GAG-CP linker biosynthetic paths, *B3GAT3* variants present overlapping overt phenotypes depending on the variant, including traits of Larsen, Antley-Bixler, Shprintzen-Goldberg, Geroderma osteodysplastica, and spEDS syndromes[46,48]. Disease's traits combined to data on substrate selectivity and B3GAT3 affinities to different core protein, agree the overlaying GAG-CP linker synthetic paths (Fig. 6A) and corroborates that linkeropathies are a phenotypic continuum bridging EDS and skeletal disorders[48]. For instance, it has been shown that phosphorylation of Xyl increases the affinity of B3GAT3 for its substrate in the common HSPG/CSPG pathway, while sulfation at C6 of Gal-II increases the affinity of the enzyme for its substrate in the small-CSPG pathway. As B3GAT3 is predominantly associated to PXYLP1 in the HSPG/CSPG path, dephosphorylation of the GlcA-Gal-Gal-Xyl(P)-(Ser/CP) linker immediately after its formation produces the substrate required for the HS chain initiation by EXTL3 (Fig. 6A and 6B). Alternatively, in the same path, a PXYLP1-free B3GAT3 fraction can also glycosylate Gal-Gal-Xyl(P)-(Ser/CP) to produce GlcA-Gal-Gal-Xyl(P)-(Ser/CP), which initiates CS elongation through the here called 'neurocan path', which was evidenced by the phosphorylated GAG-CP linker observed in neurocan[40]. In a less privileged HSPG/CSPG-derived path, GlcA-Gal-Gal-Xyl(P)-(Ser/CP) is dephosphorylated and subsequently glycosylated by CS *N*-acetyl galactosamine transferase 1 (CSGALNACT1), promoting the here called 'aggrecan path', evidenced by the non-sulfated/non-phosphorylated GAG-CP linker that prevails in aggrecan[40]. Interestingly, studies in substrate selectivity in the HSPG/CSPG indicate that the HSPG path is preferred over the 'aggrecan path'[40-42], in agreement with the predominant skeletal phenotype observed in most linkeropathies[48]. Moreover, substrate selectivity at the FAM20B and BGALT6 involving steps indicates that as the HSPG/CSPG path is favored *vs* the 'CSPG-decorin' path. This can explain the predominant connective tissue phenotype characteristic of B3GAT3-related linkeropathies. Because of the central involvement of the enzyme in normal embryonic development, *B3GAT3* was allocated to the Group I of HSPG (and CSPG) biosynthetic machinery genes (Fig. 4 and Fig. 5).

*PXYLP1* (Chr.3; ID: 92370)

Phosphoxylose phosphatase 1 (PXYLP1) is a type II transmembrane phosphatase that dephosphorylate Xyl(P) residues in the GlcA-Gal-Gal-Xyl(P)-(Ser/CP) linker tetrasaccharide before HS or CS polymerization starts (Fig. 6A). *PXYLP1* transcripts are ubiquitously expressed (Fig. 4), suggesting the importance of this gene in homeostasis. In human, *PXYLP1* pathogenic variants have not been reported for the single gene, and *Pxylp1* knockout mice have not being generated. This might be due to lethality at early embryonic stages due to a blocked HS synthesis, or to non-pathologic phenotypes (less probable). Indeed, in the Golgi, PXYLP1 interacts with either B3GAT3 or with the CSGALNACT1 to respectively promote initiation of HS or CS synthesis[49,50] (Fig. 6A). Accordingly, HS initiation by EXTL3 is blocked in the absence of PXYLP1[50], while CS elongation can still occur[49] (Fig. 6A). As abundancies of B3GAT3 and CSGALNACT1 determine their interactions with PXYLP1[49,50], the phosphatase might thus play important roles in determining whether a GAG chain will elongate in part-time proteoglycans[49]. *PXYLP1* was here included in the Group I of ubiquitously expressed HSPG biosynthetic genes required for healthy life (Fig. 4 and Fig. 5).

***Nucleotide regulators***

*CANT1* (Chr.17; ID:124583) and *SLC10A7* (Chr.4; ID: 84068)

*CANT1* codes for the ubiquitous calcium activated nucleotidase-1 (CANT1) (Fig. 4), a calcium dependent enzyme that hydrolyses uridine diphosphate nucleotide (UDP) to produce uridine monophosphate[27] and inorganic phosphate (Pi) in the ER and in the Golgi (Fig. 6A). UDP nucleotides (UDP-Xyl, UDP-Gal, UDP-GlcA, UDP-GlcNAc, etc.) are the glycosylation reaction donors that are synthesized in the cytoplasm and transported into the Golgi by bidirectional transporters[51]. In the Golgi, after its production by glycosylation reactions, UDP must be hydrolyzed by Golgi nucleotidases, including CANT1, to produce Pi and UMP. UMP is a strong inhibitor of glycosyltransferases and must then be redirected to the cytosol. Thus, by avoiding the inhibition of glycosyltransferases including XYLTs, CANT1 promotes the synthesis of the GAG-CP linker[51]. Thus, CANT1 genetic variability can be expected to affect both HS and CS paths, leading to clinical traits like those produced by variants in the genes involved in the first steps of the GAG-CP linker synthesis[52]. Accordingly, in human, *CANT1* genetic variations result in Desbuquois dysplasia (DBQD), which regroups traits observed in the XYLT1-related DBQD-2 and in B4GATT7-Larsen syndrome, including severe prenatal and postnatal growth retardation, joint laxity, short extremities, and progressive scoliosis[52]. This disease phenotype is recapitulated in *Cant1*$^{-/-}$ mice[53], suggesting the inhibition of both CSPG and HSPG/CSPG paths. Moreover, since CANT1 is calcium dependent, it can be expected that an altered calcium homeostasis, as that due to mutations in $Ca^{2+}$ transporter SLC10A7 (Fig. 6A), results in traits overlapping with those in CANT1 (DBQD) and XYLT1 (DBQD-2). Accordingly, the *Slc10a7*$^{-/-}$ mice showed skeletal dysplasia with such traits[54]. Thus, *CANT1* and *SLC10A7* were both included in the Group I of ubiquitously expressed HSPG biosynthetic machinery genes required for normal embryonic development (Fig. 4 and Fig. 5).

***Other Golgi transporters***

By compromising the levels of sugar and sulfate donors in the Golgi, genetic variability of nucleotide sugar transporters and sulfate transporters can be expected to dramatically alter GAG synthesis and consequently global tissue homeostasis[51,55]. The sulfate ion ($SO_4^{2-}$) transporter SLC26A2 (*SLC26A2*), functions as a $SO_4^{2-}$/2$OH^-$, $SO_4^{2-}$/2$Cl^-$, and $SO_4^{2-}$/$OH^-$/$Cl^-$ exchanger at the cell membrane, and supplies $SO_4^{2-}$ for the biosynthesis of the glycosaminoglycan sulfate donor 3'-phosphoadenosine-5'-phosphosulfate (PAPS) in the cytosol before its transfer to the Golgi. *SLC26A2* clinical variants can then be expected to decrease PAPS levels in the Golgi, affecting GAG from the first steps of their biosynthesis. Accordingly, *SLC26A2* clinical variants are responsible of inborn genetic skeletal disorders, which severities depend on the *SLC26A2* mutations and vary from mild recessive multiple epiphyseal dysplasia (malformations of hands, feet, and knees with possible scoliosis)[56] to perinatally lethal atelosteogenesis 2 (chondrodysplasia with dysmorphic syndrome) passing through diastrophic dysplasia (dwarfism with short legs and arms) and achondrogenesis 1B (chondrodysplasia with severe micromelia)[57]. Similarly, genetic variability in any of the transporters that assure UDP-sugars levels in the Golgi, as *SLC35A* (UDP-GlcNAc and UDP-Gal transporter) and *SCL35A3* (UDP-GlcNAc transporter) (Fig. 6), can affect the biosynthesis of GAGs and of several other glycosylated molecules depending on the tissue n where they are expressed. For instance, *SCL35A2* (UDP-GlcNAc transporter) variants can cause congenital disorders with CNS-related symptoms such as epilepsy and autism[58].

**Group I – Essential wildly expressed HS chain biosynthetic genes**

Elongation and maturation of a whole HS chain requires the action of glycosyl transferases, sulfotransferases, and epimerase. Without a priori and based on the phenotypes of their clinical variants and profiles of expression (Fig. 4), *EXTL3*, *EXT1*, *EXT2*, *GLCE*, *NDST1*, *HS2ST1*, and *HS6ST1* were included in the Group I of HSPG biosynthetic machinery genes. Interestingly, these ubiquitously expressed genes can by themselves assure the synthesis of a complete HS chain by assuring chain initiation, elongation, epimerization, and sulfation at relevant positions (*N*, 2-*O*- and 6-*O*-sulfation). It is then possible that, because of their ubiquitous expression, these genes assure production of ‘ubiquitous HS chains’ (uHS) carrying the sulfation patterns dictated by NDST1, HS2ST1, and HS6ST21 substrate specificities, as detailed below.

***Heparan chain initiation***

*EXTL3* (Chr.8; ID:2137)

The *Exostosin like glycosyltransferase 3* (*EXTL3*) gene encodes the type II integral membrane protein EXTL3 that catalyzes the transfer of a GlcNAc residue from UDP-GlcNAc to the non-reducing end of the GAG-CP linker tetrasaccharide GlcA-Gal-Gal-Xyl-(Ser/CP) to form GlcNAc-GlcA-Gal-Gal-Xyl-(Ser/CP), which initiates HS biosynthesis[59,60] (Fig. 6A and 6B). EXTL3 is the only known initiator of HS chain in all organisms, and it has been suggested that its alternative splicing contribute to the regulation of HS *vs* CS levels[60,61]. Indeed, in case of defective EXTL3 activity or when the enzyme is mis-localized due to genetic variability, GlcA-Gal-Gal-Xyl-(Ser/CP) accumulates [and possibly GlcA-Gal-Gal-Xyl(P)-(Ser/CP) in severe variants], leading to CS elongation[49,60,61] (Fig. 6A). Accordingly, Human *EXTL3* genetic variability results in decreased HS synthesis with increased CS production[60]. Clinical phenotypes associated to *EXTL3* mutations include skeletal dysplasia, developmental delay, severe epispondylo-metaphyseal dysplasia with immunodeficiency, and in most severe cases neuromotor and brain development delay[60]. In accord with the fundamental role of EXTL3 in life, *Extl3*$^{-/-}$ mice fail to produce HS chains and die during early embryonic development[60,62]. Therefore, *EXTL3* was allocated to the Group I of ubiquitous HSPG biosynthetic genes essential for normal embryonic development and homeostasis (Fig. 4 and Fig. 5).

***Heparan chain elongation***

*EXT1* (Chr.8; ID: 2131) and *EXT2* (Chr.11; ID: 2132)

*Exostosin glycosyltransferase 1 (EXT1)* and *exostosin glycosyltransferase 2* (*EXT2*) code for the ubiquitous type II integral membrane proteins EXT1 and EXT2, responsible of HS chain elongation in all tissues (Fig. 2). These enzymes act as a hetero-oligomeric complex that alternatingly transfer GlcA and GlcNAc to the growing HS chain. Both EXT1 and EXT2 are required for HS chain elongation and neither of the two proteins can substitute the other one. Indeed, it is likely that, while EXT1 assures the glycosyl transferase activity, EXT2 assists by assuring correct EXT1 folding and Golgian location[59,63]. The essential role of EXT1 and EXT2 in normal development was shown in *Ext1/Ext2* deficient embryos, which failed to gastrulate and died in utero by embryonic day 7.5. However, compound heterozygous *Ext1*$^{+/-}$;*Ext2*$^{+/-}$ mice were viable but developed multiple osteochondromas by 2 months of age[64,65]. Although chondrocytic defects were observed in that mice, several other defects also occur, as shown by the *Ext1* specific deletion in epithelial cells during lung development, which resulted in reduced branching number, expanded branching tips, and several mesenchymal developmental defects[66]. Indeed, defective EXT1/EXT2 leads to accumulation of the GlcNAc-GlcA-Gal-Gal-Xyl-(Ser/CP) substrate, which redirects the GAG synthesis through the CS pathway, enriching CS-aggrecan production (Fig. 6) and thus resulting in an increased CS/HS ratio, which results in an exacerbated differentiation of perichondrial progenitors into chondrocytes[63]. Accordingly, in human, homozygous or compound heterozygous mutations in *EXT1* and/or *EXT2* are causative of hereditary multiple exostoses, a dominantly inherited genetic disorder characterized by the formation of cartilage-capped bony outgrowths at the epiphyseal growth plates, with a variable clinical phenotype depending on the variant[63]. Interestingly, *EXT1*-related alterations of HS levels in tumor cells has been shown to participates to the progression of many cancers[67]. Because of its severe clinical phenotypes and wide expression, *EXT1* and *EXTL2* were placed in the Group I of ubiquitous HSPG biosynthetic genes required for normal embryonic development and tissue homeostasis (Fig. 4 and Fig. 5).

***HS chain maturation***

*NDST1* (Chr.5; ID: 3340)

N-*deacetylase/*N-*sulfotransferase 1* (*NDST1*) codes for the widely expressed NDST1 enzyme that resides in the Golgi membrane and presents multiple isoforms resultant from alternative splicing[68]. Among all *NDSTs*, only *NDST1* shows a ubiquitous expression (Fig. 4). NDST1 produces GlcNS residues by deacetylating GlcNAc and transferring a sulfate group form PAPS to GlcN. The enzyme can initiate *N*-sulfation at any internal GlcNAc residue, and sulfation proceeds from the nonreducing end toward the reducing end, to stop at least five saccharide units away from the GAG-CP linker or from previously existing GlcNS residues; this generates intermediary NA domains that separate GlcNS clusters[69]. Interestingly, it has been stablished that the length of the *N*-sulfated clusters made by NDST1 is much shorter than that made by NDST2[69,70]. In human, non-null homozygous *NDST1* mutations are known, in agreement with the phenotype of *Ndst1*$^{-/-}$ mice, which perinatally die with immature lungs, delayed mineralization of the skeleton, vascular defects, craniofacial dysplasia, eye developmental defects, etc.[71]. Indeed, these mice produce HS with under sulfated structures, indicating that other NDSTs cannot compensate NDST1 function when the enzyme is completely missing. However, the possibility to live with partially active NDST1 is possible, as suggested by the varying pathologic phenotypes resultant from compound heterozygous *NDST1* mutations in human. For instance, combined heterozygous *NDST1* mutations have been associated to developmental delays, intellectual disability, muscular hypotonia, ataxia, history of a seizures, epilepsy, gastroesophageal reflux, and minor malformations[72,73]. These predominant brain and muscle phenotypes can be explained by a partial compensation by NDST2 in most organs, but not in brain and skeletal muscle, in where NDST2 is absent or undetectable during adulthood (Fig. 4). For all these reasons, *NDST1* was allocated into the Group I of ubiquitous HSPG biosynthetic genes required for normal embryonic development (Fig. 4 and Fig. 5).

*GLCE* (Chr.15; ID: 26035)

D-*glucuronyl C5-epimerase (GLCE)* codes for C5-epimerase, the widely expressed type II integral membrane enzyme that epimerizes C5 in GlcA to form IdoA, which favors HS2ST recognition and confers to HS chains the flexibility required for their interaction with HBP ligands[74]. C5-epimerase acts under a substrate control dictated by the distribution of GlcNS and GlcNAc residues flanking (+1 site) the GlcA that will be converted into IdoA[75,76]. Enzyme kinetics have shown that C5-epimerisation can readily occur in GlcA residues flanked by GlcNS (-GlcNS-**GlcA**-GlcNS/GlcNAc-) to reversibly form IdoA (-GlcNS-**IdoA**-GlcNS/GlcNAc-), while presence of a GlcNAc residue at the non-reducing neighboring side (-GlcNAc-**IdoA**-GlcNS-**IdoA**-GlcNS/GlcNAc-) or of 2-*O*-sulfated IdoA (-GlcNS-**IdoA2S**-GlcNS-**IdoA**-GlcNS/GlcNAc-), avoids reaction reversibility and stabilizes the epimerized product[75,76]. Indeed, this conversion occurs under a stringent substrate specificity in which 2-*O*-sulfation of IdoA, and/or 6-*O*- or 3-*O*-sulfation of GlcN units at subsite −2 to +2, leads to nonproductive substrate binding[75]. Similarly, 6-*O*-sulfation in -GlcNS6S-**GlcA**-GlcNS6S- was shown to inhibit C-5 epimerase activity[75,76]. In the other side, the association of C-5 epimerase and HS2ST, which assures 2-*O*-sulfation, allows the irreversible and thus stable generation of contiguous epimerized and 2-*O*-sulfated domains in the HS chain[77] (Fig. 6B). In agreement with an essential role in normal embryonic development, neither single *GLCE* homozygous null mutations nor heterozygote compound pathogenic variants are known in human, possible because lethal, as supported by the *Glce*$^{-/-}$ mice, which neonatally die with severe developmental abnormalities affecting several organs including kidney, lung, skeleton, spleen, thymus, lymph node, etc.[74]. However, SNP has been associated with hypertension and cerebrovascular events[74,78]. The importance of *GLCE* in the regulation of cell behavior is also suggested by its potential tumor suppressor activity, for instance in breast and lung carcinogenesis[79]. Because of its requirement for normal embryonic development and physiology, *GLCE* was placed in the Group I of ubiquitous HSPG biosynthetic genes essential for life (Fig. 4 and Fig. 5).

*HS2ST1* (Chr.1; ID: 9653)

*Heparan sulfate 2*-O-*sulfotransferase 1* (*HS2ST1*) codes for the ubiquitously expressed HS2ST type II integral membrane protein (Fig. 4), which transfers a sulfate group form PAPS to the C2 position of hexuronic acid residues in HS, with preference for IdoA over GlcA as a substrate[80,81]. In the Golgi, HS2ST can interact with GLCE in a close functional relationship, which centrally contributes to the generation of IdoA2S residues in the HS chain (Fig. 6B). In human, *HS2ST1* clinical variants are unknown, possibly since not viable, as suggested by the *Hs2st1*$^{-/-}$ mouse, which died at the neonatal period with developmental failure and an onset of abnormalities in several organs including kidney, which completely failed after mid-gestation[81]. Interestingly, although HS in organs of the *Hs2st1*$^{-/-}$ mice lacked 2-*O*-sulfation, the overall HS charge density was maintained through increased *N*- and 6-*O*-

sulfations. However this did not rescue the mice developmental failure, in agreement with the importance of specific 2-*O*-sulfation in normal embryonic development. Interestingly, *HS2ST1* expression increases in adult tissues after injury, probably because the enzyme action favors the recovery of tissue homeostasis[82]. Thus, since HS2ST is a widely expressed and required for the synthesis of functional HS chains, *HS2ST1* was allocated into the Group I of ubiquitous HSPG biosynthetic genes required for normal embryonic development (Fig. 4 and Fig. 5).

*HS6ST1* (Chr.2; ID: 9394)

*Heparan sulfate 6-O-sulfotransferase 1 (HS6ST1)* codes for the ubiquitously expressed HS6ST1, a type II integral membrane protein that transfers sulfate group from PAPS to C6 of GlcN in HexA-GlcNAc/NS residues to generate HexA-GlcNAc/NS**6S** along, and at the reducing extremities, the HS chain[83] (Fig. 6B). As 6-*O*-sulfation in -GlcNS**6S**-GlcA-GlcNS**6S**- inhibits C-5 epimerase activity on the GlcA motif[75,76], HS6ST1 favors formation of HS NA/NS domains, which characteristically display low content of IdoA2S residues. In human, *HS6ST1* clinical variants are unknown, possibly because carriers are not viable, as suggested by the *Hs6st1*$^{-/-}$ mice, which failed to thrive or died at late embryonic and perinatal stages with growth retardation, lung defects, and abnormal angiogenesis, among other abnormalities except in kidney[83,84]. Interestingly, the lack of noticeable abnormalities in the kidney of these mice suggests that HS6ST2, which is expressed in kidney at relatively high levels (Fig. 4), might compensate HS6ST1 lack[83,84], as recently suggested in mice with *Hs6st1* systemic inactivation[85]. Moreover, *HS6ST1* SNPs have been associated to risk of delayed puberty and to idiopathic hypogonadotrophic hypogonadism[86], and HS6ST1 and HS6ST2 were both proposed as candidate targets for ovarian failure during menopause[86]. Irrespective to genetic variability, a critical role of *HS6ST1* is also suggested by a high *HS6ST1* mRNA observed in renal fibrosis associated with chronic rejection[87], idiopathic pulmonary fibrosis[88], and osteoarthritic cartilage[89]. This suggest that while HS made by HS6ST1 are involved in the maintain of tissue homeostasis, they might not be suitable for an appropriate tissue response to injury, although this is speculative and remains to be investigated. Because of its ubiquitous tissue expression and critical role in normal embryonic development, *HS6ST1* was placed in the Group I of HSPG biosynthetic genes (Fig. 4 and Fig. 5).

**Group I – Ubiquitous HS chain remodeling machinery genes**

*SULF2* (Chr.20; ID: 55959)

*Sulfatase 2* (*SULF2*) encodes the broadly expressed extracellular HS endosulfatase 2 that is secreted to the extracellular matrix or localized in lipid rafts at the cell surface[90]. SULF2 selectively removes 6-*O*-sulfate groups from HS in the extracellular space starting from the non-reducing end through the reducing end along highly sulfated NS domains in HS chains. Interestingly, by desulfating HS, SULF2 favors tumor growth, likely by enhancing cell signaling[90]. The importance of *SULF2* in normal tissue homeostasis was demonstrated in the *Sulf2*$^{-/-}$ mouse, which showed partial embryonic lethality (~35% of mice) with developmental malformations particularly affecting the nervous system[91]. However, the double knockout *Sulf1*$^{-/-}$/*Sulf2*$^{-/-}$ showed to be severely lethal, whereas the *Sulf1*-deficient mouse showed only mild phenotype. It remains to be clarified whether the partial lethality in *Sulf2*$^{-/-}$ mice was associated to *Sulf1* compensatory mechanisms, functional co-operativity, limited redundancy, or all[92]. Interestingly, in human, genetic variations in *SULF2* have been reported as risk factors for altered regulation of lipoprotein metabolism[93] and therefore it has been proposed as a potential therapeutic target for atherogenic dyslipidemia in obesity and type 2 diabetes mellitus[94,95]. Because of the lethality detected in *Sulf2*$^{-/-}$ mice and for its ubiquitous expression, *SULF2* was placed in the Group I of HSPG biosynthetic machinery genes (Fig. 4 and Fig. 5).

#### Group II. Essential tissue restricted HSPG biosynthetic genes

##### Group II – Essential tissue restricted HS remodeling control

*HPSE2* (Chr.10; ID: 60495)

*Heparanase 2* (*HPSE2*) codes for the enzymatically inactive heparanase 2 that shows tissue restricted expression (Fig. 4) and close homology (40% ) with the enzymatically active heparanase [96]. Although heparanase 2 has no enzymatic activity, the protein displays higher affinity for HS than the enzymatically active heparanase and can function as its inhibitor [96]. Under basal physiologic conditions, HPSE2 is predominant expressed in the urinary track, followed by prostate and endometrium, with some expression in other tissues in which response to stimuli is central for proper function, including gastrointestinal and reproductive systems (Fig. 4). In human, biallelic mutations in the *HPSE2* gene have been associated with Urofacial or Ochoa Syndrome (UFS or UFOS), characterized by incomplete urinary bladder emptying and a facial grimace upon smiling, and which is also caused by mutation sin the LRGI2 gene, which encodes leucine-rich-repeats and immunoglobulin-like-domains 2 and modulates growth factor signaling, [97,98]. However, in up to 16% of UFS or UFOS patients, no HPSE2 or LRGI2 associated mutations have been found [97], suggesting that *HPSE2* SNP contributes to a more complex polygenic variabilities leading to the altered condition. Accordingly, there is high clinical variability among patients with UFS and not all present HPSE2 and LRGI2 significant variability. The involvement of HPSE2 in larger phenomena is supported by genetic variability associations with presence of nocturnal lagophthalmos in some patients[96] and with the separation of migraine with and without depression[99]. The essentialness of *Hpse2* in response to physiologic stimuli was confirmed in homozygous *Hpse2* gene trap mouse, which developed normally but showed deficient myogenic and neurogenic responses to chemical and electrical field stimulation.

Accordingly, mutant mice failed to gain weight after weaning and die when maintained for more than 1 month of age [100]. *HPSE2* is the only gene allocated to the Group II of HSPG biosynthetic machinery genes (Fig. 4 and Fig. 5).

#### Group III. Non-essential widely expressed HSPG biosynthetic genes

##### Group III – Non-essential widely expressed HSPG core proteins

*GPC1* (Chr.2; ID: 2817)

*GPC1* encodes the widely expressed glypican 1 core protein (Fig. 4). Absence of pathogenic mutations in this gene agrees with the phenotype of the *Gpc1*$^{-/-}$ mouse, which develops normally, is viable and fertile[101], although showed subtle alterations on brain patterning as those observed in *GPC3*$^{-/-}$mice[102]. The involvement of *GPC1* in neurologic adaptative behavior is suggested by role *GPC1* polymorphism correlation with risk to schizophrenia when using brain activation as a quantitative phenotype [103]. Variability in *GPC1* has also been observed in people susceptible to biliary atresia susceptibility[104]. Besides a possible role in the regulation of tissue homeostasis, which remains to be established, glypican 1 was also involved in growth factor signaling in several cancer cells[105], including in pancreatic, prostate, breast and cervical cancers[17]. Because its wide expression and non-requirement for normal embryonic development, *GPC1* was allocated to the Group III of HSPG biosynthetic machinery genes (Fig. 4 and Fig. 5).

*Syndecans*

The *syndecan* (*SDC*) family of genes, which includes *SDC1*, *SDC2*, *SDC3*, and *SDC4*, codes for type I transmembrane proteins cores formed by an ectodomain, a transmembrane region and an intracellular domain. All syndecans have been involved in the regulation of cell-matrix interactions and in cell signaling[106]. Moreover, syndecans can act as endocytosis receptors and are also involved in the uptake of exosomes. Interestingly, the intracellular C-terminus in the four syndecans harbors a unique EFYA sequence that binds to the PDZ-containing proteins, which contribute to the anchoring of transmembrane proteins to the cytoskeleton, holding together large protein complexes along the cell membrane[107]. In the extracellular space, all syndecans can be shed into the pericellular environment through the action of matrix metalloproteinases (MMPs). Although ubiquitously expressed, any pathogenic *SDC* clinical variant has been reported in human and the available knockout mice develop normally, are viable, fertile, and lives normally, signifying a non-essential role for development and homeostasis. However, syndecans have shown to play important roles during tissue response to physiological or pathological stimuli, as in wound healing, inflammation, host defense to instigating agents, and during tumor cell growth and invasiveness[108]. Because their wide expression and non-requirement for normal embryonic development and homeostasis, the four *SDCs* were included in the Group III of HSPG biosynthetic machinery genes.

*SDC1* (Chr.2; ID: 6382)

*SDC1* codes for syndecan 1, a widely expressed HSPG core protein that shows predominant expression in tissues exposed to external stimuli, as the gastrointestinal track, skin, liver, etc. (Fig. 4). In human, non-clinical variants are known for *SDC1*, in agreement with the non-deleterious phenotype observed in the *Sdc1*$^{-/-}$ mouse, which develops, lives and reproduces normally[109]. However, SDC1 genetic variables have been associated to increase susceptibility to multiple sclerosis[110] and with severity and vulnerability of coronary plaque in patients with coronary artery disease[111]. In agreement with a possible role in the regulation of biological events related to stimuli, the *Sdc1*$^{-/-}$ mouse displays altered capacities to recover when challenged with instigating agents, as microbial infection, or ischemia, suggesting its involvement in adaptative functions. *In vitro*, syndecan 1 regulates cell adhesion, cell migration, endocytosis, exosome formation, etc.[112]. These processes are central to tissue defense to aggression and to the control of fibrosis, inflammation, and re-vascularization[113,114]. Accordingly, *SDC1* altered expression often impacts the clinical course of cancers, as evidenced by its prognostic significance[113]. An emerging new role for shed syndecan-1 is related to its ability to reach the nuclei in a variety of cells, including tumor cells[115]. Because its wide expression and non-requirement for normal embryonic development and physiology, and for its involvement in tissue adaptative responses to injury or disease instigating conditions, *SDC1* was allocated to the Group III of HSPG biosynthetic machinery genes (Fig. 4 and Fig. 5).

*SDC2* (Chr.8; ID: 6383)

Syndecan 2 core protein *is coded by SDC2*, which is widely expressed (Fig. 4). Although *Sdc2*$^{-/-}$ mice have not been generated, the absence of reports on pathogenic genetic variants in human suggests a non-essential role in normal development and physiology. However, a GWAS has shown association to risk of posttraumatic stress disorder in a cohort of Iraq-Afghanistan era veterans [116]. Moreover, syndecan 2 has been involved in the processes of fibrosis[117], inflammation[118], and cancer progression[119]. This might be related to the syndecan 2 role that assures connections between the cell cytoskeleton and the extracellular environment, particularly at the epithelial-mesenchymal interface[119], and during formation of specialized membrane domains requiring redefinition during adaptative responses or during tissue recovery[120]. Similarly, in brain, syndecan 2 is involved in the continuous redefinition of specialized membrane domains in dendritic filopodia and dendritic spines in responsive neurons[121,122]. Because its wide expression, its non-requirement for normal embryonic development and physiology, and its central involvement in tissue or cell responsiveness, *SDC2* was allocated to the Group III of HSPG biosynthetic machinery genes (Fig. 4 and Fig. 5).

*SDC3* (Chr.1; ID: 9672)

*SDC3* is a ubiquitously expressed gene that shows some predominant expression in the immune and adrenal systems and in brain (Fig. 4). Although no *SDC3* pathogenic variants are known, *SDC3* polymorphism has been associated to a reduced risk to obesity[123], positive association have been found with metabolic syndrome[124], with body composition in patients with schizophrenia under treatment with antipsychotic drugs[125], and with female hyperandrogenism[126]. Accordingly, the *Sdc3*$^{-/-}$ mouse develops normally, is viable and fertile, but exhibits altered feeding behavior with resistance to obesity[127]. This suggests that syndecan 3 might be involved in fine events related to different behavioral response, as satiety, and possibly to other responses in tissues in which the gene is expressed. Besides, *SDC3* is part of the clustering of genes involved in cancer progression[128]. Because its wide expression with non-requirement for normal embryonic development, *SDC3* was allocated to the Group III of HSPG biosynthetic machinery genes (Fig. 4 and Fig. 5).

*SDC4* (Chr.20; ID: 6385)

*SDC4* is a ubiquitously expressed gene and is the major syndecan expressed in most tissues (Fig. 4). To date, *SDC4* clinical variants have not yet been reported, in agreement with phenotype of the *Sdc4*$^{-/-}$ mouse, which is viable, fertile, and macroscopically indistinguishable from wild-type littermates[129]. However, polymorphism has been associated with regulation of whole-body energy metabolism[130] and with longevity and lipid profile in healthy elderly[131]. As other syndecans, *SDC4* expression contributes to tissue response to stimuli or trauma, as observed during wound healing in *Sdc4*$^{-/-}$ mice, which showed statistically significant delayed healing of skin wounds and impaired angiogenesis[129]. Indeed, as other syndecans, syndecan 4 can mediate numerous cellular processes through signaling pathways that affect cellular proliferation, migration, mechano-transduction, and endocytosis[108]. Because its wide expression and its non-requirement for normal embryonic development, but required for proper response to stimuli or trauma, *SDC4* was allocated to the Group III of HSPG biosynthetic machinery genes (Fig. 4 and Fig. 5).

**Group III – Non-essential widely expressed HS chain remodeling genes**

*EXTL2* (Chr.1; ID: 2135)

The widely expressed *EXTL2* gene codes for the exostosin like glycosyltransferase 2 (EXTL2) enzyme that acts as a ‘quality control’ when the proteoglycan biosynthetic machinery is over-activated during loss of tissue homeostasis[132]. EXTL2 specifically transfers a GlcNAc residue to the GlcA-Gal-Gal-Xyl(P)-Ser/CP tetrasaccharide to generate GlcNAc-GlcA-Gal-Gal-Xyl(P)-Ser/CP, which blocks both HS and CS biosynthesis. *Extl2* knockout mice develops normally, are viable and live healthy and non *EXTL2* clinical variants have been reported in human. However, after tissue insult, EXTL2 favors tissue recovery by avoiding excessive CS and HS biosynthesis, as shown by impaired tissue regeneration in mice models of liver and kidney failure in which *Extl2* was knocked out[133,134]. Because EXTL2 is not essential for normal embryonic development, and since its activity is relevant when a tissue needs to recover, *EXTL2* was allocated to the Group III of HSPG biosynthetic machinery genes (Fig. 4 and Fig. 5).

*SULF1* (Chr.8; ID: 23213)

*Sulfatase 1* (*SULF1*) encodes the broadly expressed extracellular HS endosulfatase-1, which selectively removes 6-*O*-sulfate groups from HS. SULF1 is secreted through the Golgi to be subsequently localized at the cell surface[90]. Interestingly, *Sulf1*$^{-/-}$ mice show non-overt phenotypes, whereas *Sulf2*$^{-/-}$ mice show partial lethality and *Sulf1*$^{-/-}$/*Sulf2*$^{-/-}$ show severe lethality, suggesting that SULF1 can be compensated by SULF2[91]. In human, SULF1 gene polymorphisms have been associated to increased risk to multiple sclerosis[110], fetus failure in IVF technique[135], and Preeclampsia[136]. SULF1 has been involved in events in which tissue homeostasis is lost[90,94,137], as inflammation, wound healing, and cancer. Interestingly, in these processes SULF1 protects possibly by avoiding excessive accumulation of the ubiquitous NA/NS domains in HS that promote cell signaling. On the opposite, SULF2 is associated with decreased survival[90], possibly by the abnormal accumulation of NA/NS-like HS domains resulting from 6-*O*-desulfation of NS domains. Since *SULF1* is widely expressed and *Sulf1*$^{-/-}$ mice are viable and evolve healthy, unless homeostasis is lost, this gene was placed in the Group III of genes involved in the maintain of tissue homeostasis, particularly favoring tissue recovery when homeostasis is loss. *SULF1* was allocated to the Group III of HSPG biosynthetic machinery genes (Fig. 4 and Fig. 5).

*HPSE* (Chr.4; ID: 10855) and HPSE2 (Chr.10; ID: 60495)

Heparanase (HPSE) is the only enzyme in mammals known to exert glycolytic cleavage of HS chains in the extracellular space[138]. *HPSE* is ubiquitously expressed (Fig. 4) and several transcript variants encoding different isoforms have been found for this gene. Interestingly, its activity can be regulated by its inactive close homolog *heparanase-2* (*HPSE2*), which shows tissue restricted expression (Fig. 4). HPSE cleaves β(1,4)-glycosidic bonds between GlcN residues and GlcA at a limited number of sites in HS[139]. This participates to the remodeling of extracellular matrices and basement membranes, allowing cell movement and release of HBP (cytokines, growth factors and several other biological signals) from HS scaffolds in the extracellular space[138]. Although widely distributed, HPSE exhibits only very low levels of expression, possibly because of its principal role during tissue remodeling processes, as wound healing, angiogenesis, inflammation. Accordingly, *Hpse*$^{-/-}$ mice develop normally, are viable, anatomically normal, and fertile[140] and non-pathologic variants for this gene are known in human. However, gene variability has been found to increase risk of graft-versus-host disease, which is a major complication following allogeneic hematopoietic cell transplantation (HCT) and occurs when donor T cells respond to histo-incompatible antigens on the host tissues[141,142]. In agreement with a role in matrix remodeling, HPSE is overexpressed in pathological conditions where active matrix remodeling is highly active, such as fibrosis, inflammation, and angiogenesis, and in a variety of pathologies including kidney diseases, bone osteolysis, preeclamptic placenta, multiple sclerosis, autoimmune disease, viral infection, thrombosis, atherosclerosis, sepsis, diabetes, cancer progression and metastasis[143]. Since HPSE is widely expressed and the *Hpse*$^{-/-}$ mouse is viable and fertile, *HPSE* was allocated to the Group III of HSPG biosynthetic machinery genes (Fig. 4 and Fig. 5).

#### Group IV. Non-essential tissue restricted HSPG biosynthetic genes

##### Group IV – Non-essential tissue restricted HSPG core proteins

*GPC2* (Chr.20; ID: 221914)

*GPC2* encodes the glypican 2 core protein, which is predominantly expressed in brain and in some reproductive organs, particularly in endometrium and testis, although the gene is also expressed in other tissues but at low extent (Fig. 4). In human, *GPC2* clinical variants have not been reported to this date, and the *Gpc2* knockout mouse has not yet been generated. However, *GPC2* polymorphism is part of the polygenic scores for risk of developing Alzheimer's disease[144]. Moreover, some data suggest that *GPC2* expression is require for the adaptative responses of tissues in which it is expressed[145]. For instance, in brain, both *GPC2* expression and glypican 2 protein levels increase when neurogenesis is stimulated in the hippocampus of the adult brain and decrease when neurogenesis is selectively ablated in transgenic models[145]. Accordingly, glypican 2 was proposed as a potential marker to monitor adult neurogenesis in cerebrospinal fluid[145]. Interestingly, *GPC2* has been correlated with poor survival in neuroblastoma[66]. Because its tissue restricted expression, and although its role in reproductive organs is still unknown, *GPC2* was included in the Group IV of HSPG biosynthetic genes (Fig. 4 and Fig. 5).

*GPC5* (Chr.13; ID: 2262)

Glypican 5 is a HSPG core protein coded by the *GPC5* gene, which expression is restricted to only some tissues in the adult human body, particularly kidney and testis (Fig. 4). Although there is not *GPC5* clinical variants in human and the *Gpc5* knockout mice has not yet been generated, genetic variants in association with other genes variants, have been associated to vulnerability to acquired nephrotic syndrome[146], multiple sclerosis[147], autoimmune thyroid disease[148]. Moreover, *GPC5* seems to be required for the adaptative responses to injury in the tissues in which it is expressed. For instance, *GPC5* was associated to podocyte response to injury and enhanced glypican 5 urinary levels were associated to nephropathy progression in type 2 diabetes patients[33]. Besides, *SDC5* has been proposed as a tumor suppressor in lung and prostate cancer[149,150]. Because its tissue restricted expression, with possible requirement for response to injury, *GPC5* was included in the Group IV of HSPG biosynthetic machinery genes (Fig. 4 and Fig. 5).

**Group IV – Non-essential tissue restricted specialized HS chain maturation**

*NDST2* (Chr.10; ID: 8509)

*NDST2,* a gene expressed in several but not all tissues (Fig. 4), codes for the Golgian NDST2 enzyme, which exhibits dual *N*-deacetylase and *N*-sulfotransferase activities and is involved in the production of the large NS domains in HS chains[59,68]. Under physiological conditions, NDST2 is not active in the presence of the ubiquitous NDST1, likely because in the presence of NDST1, NDST2 remains distant from the GAGosome and thus from its substrate[68,151]. Accordingly, *Ndst2*$^{-/-}$ mice are viable, fertile and show normal HS structures in all tissues, with exception of mast cells, in which NDST2 is predominantly expressed and heparin is synthetized [151]. In agreement with a non-essential role during normal development, no pathogenic *NDST2* variants are known in human. However, NDST2 SNP was found potentially associated to risk of coronary artery and chronic kidney diseases[152]. Interestingly, during tissue injury or during inflammation NDST2 becomes active[153], possibly because under stimuli the enzyme is integrated into the GAGosome, resulting in the production of longer sulfated domains carrying characteristic NS structures[151]. Although more research is required to determine how injury, inflammation, or other stimuli can favor NDST2 expression and function, *NDST2* was allocated to the Groupe IV of HS biosynthetic machinery genes (Fig. 4 and Fig. 5).

*NDST3* (Chr.4; ID: 9348) and *NDST4* (Chr.4; ID: 64579)

*N*-deacetyl, *N*-sulfotransferases 3 and 4 (NDST3 and NDST4) are both type II transmembrane enzymes that show highly restricted tissue expression, but that are particularly present in brain and spleen (Fig. 4), two organs highly receptive to immunological or behavioral stimuli, and/or, although at lower extent in other organs receptive to external stimuli, as stomach. NDST3 and NDST4 enzymes regulate HS fine structures with opposite actions, while NDST3 exhibits high deacetylase and weak sulfotransferase activity (leading to GlcNH$_2$ carrying HS chains), NDST4 shows weak deacetylase with high sulfotransferase activity (possibly to resulfate GlcNH$_2$). This might allow a molecular switch in high responsive tissues, as brain and spleen[59]. In agreement with its restricted expression and non-direct involvement in central processes required for normal embryonic development and physiology, the *Ndst3*$^{-/-}$ mouse develops normally, is fertile, and shows only subtle behavioral anomalies, with reduced anxiety and a slight hematological phenotype[154]. In the other side, the *Ndst4*$^{-/-}$ mouse exhibited no overt pathological phenotypes, although some anomalies were observed in histology images of the proximal colon in where the enzyme is not normally expressed; these images showed an increased number of goblet cells and a decreased number of colonocytes, in agreement with the

*NDST4* proposed role in colorectal tumor growth[155]. In human, non-pathogenic variants in *NDST3* or *NDST4* are known, but, in accord with their predominant expression in brain, *NDST3* genetic variants have been associated with schizophrenia and bipolar disorders[156,157], and variants in *NDST4* have been associated with reading disability and language impairment[158]. However, confirmation of the involvement of NDST3 and NDST4 in these pathological conditions requires extensive research. Thus, NDST3 and NDST4 are not essential for normal embryonic development and homeostasis, but their lack in mice results in a slight cognitive and hematological phenotype and genetic variant might contribute to disease vulnerability. Because of their tissue restricted expression, *NDST3* and *NDST4* were included in the Group IV of HSPG biosynthetic machinery genes (Fig. 4 and Fig. 5).

*HS6ST2* (Chr.X; ID: 90161)

*HS6ST2*, which codes for HS 6-*O*-sulfotransferase 2, depicts two alternative splicing variants in human, a full-length variant (HS6ST2) and a short length variant with 40 deleted amino acids (HS6ST2S). The major longer HS6ST2 variant preferentially introduces sulfate groups at C6 of GlcNS motifs adjacent to IdoA2S (IdoA2S-GlcNS), generating the sulfate rich IdoA2S-GlcNS6S disaccharide present in HS NS domain[84,159-161]. In the other side, the minor shorter variant shows no substrate specificity and can participate to the synthesis of both NS and NA/NS domains[159-161]. The partially restricted expression of *HS6ST2* in tissues suggests specialized roles, particularly in kidney and in reproductive organs, as ovary, placenta, and endometrium, in which it is predominantly expressed (Fig. 4). Although the *Hs6st2* knockout mouse has not been generated, non-severe *HS6ST2* pathogenic variants are known in human. However, genetic variability has been reported as risk factor for intellectual disability linked to chromosome X[162], in where the *HS6ST2* is located, suggesting that the NS domains produced by the enzyme can be involved in the regulation of discrete physiologic events enhanced fibroblast growth factor-mediated chondrocyte growth and differentiation[163], while HS6ST2 downregulation by miR-23-3p enhanced matrix degradation in osteoarthritis[164], in which HS6ST1 expression is enhanced[89]. This supports different biological roles of the two HS6STs in homeostasis and during pathology. Interestingly, HS6ST2 is found in body fluids, although the reason, mechanisms, and consequences of HS6ST2 secretion remains to be elucidated. Moreover, *HS6ST2* overexpression has been associated with poor prognosis in patients with gastric cancer[165]. Because of its partial tissue restricted expression, this gene was included in the Group IV of HSPG biosynthetic machinery genes (Fig. 4 and Fig. 5).

*HS6ST3* (Chr.13; ID: 266722)

*HS6ST3* codes for the tissue restricted HS 6-*O*-sulfotransferase 3 (HS6ST3), which expression is predominant in brain, although also found expressed at lower extent in some other tissues including the urinary blade (Fig. 4). Interestingly, HS6ST3 shows non substrate selectivity, as it acts on HexA-

GlcNAc/S residues to generate HexA-GlcNAc/S6S[160]. Although the *Hs6st3* knockout mouse has not yet been generated, the absence of pathogenic variants in human suggests that the gene is not essential for embryonic development and physiology. However, the importance of discrete roles of HS6ST3 in the maintain of tissue homeostasis was suggested by GWAS associating polymorphism in loci near *HS6ST3* with risk to develop obesity[166], diabetic retinopathy[167]. Moreover, HS6ST3 has also been involved in the growth and progression of breast cancer cells[168]. Since *HS6ST3* shows high tissue restricted expression and non-clinical variants are known in human, this gene was allocated to the Group IV of HSPG biosynthetic machinery genes (Fig. 4 and Fig. 5).

*HS3ST1* (Chr.4; ID: 9957)

*HS3ST1* codes for the HS glucosamine 3-*O*-sulfotransferase 1 (HS3ST1), an enzyme that introduces a sulfate group from the sulfate donor PAPS into C3 of GlcN in GlcA-GlcNS±6S to generate GlcA-GlcNS**3S**±6S in HS chains. The enzyme tolerates IdoA, but 2-*O*-sulfation specifically prevents its action[169]. HS3ST1 shows partial tissue distribution (Fig. 4), with transcripts predominantly expressed in urinary bladder, ovary, stomach, and in mastocytes, in where it participates to the production of the endogenous heparin[170]. The addition of the 3-*O*-sulfate groups to GlcN units is a relatively rare modification, present in only a limited number of chains of HS in the body, except for heparin, which is a richly 3-*O*-sulfated HS (3S-HS) used in clinics as anticoagulant and mainly extracted from porcine, bovine, or ovine intestinal mucosa[171]. As the heparin anticoagulant activity is due to its high affinity for antithrombin, anticoagulant 3S-HS are called $HS^{AT}$[171]. However, although the *Hs3st1*$^{-/-}$ mouse shows decreased $HS^{AT}$ levels, this mouse is entirely viable and fertile and its coagulation is normal, although it showed very light intrauterine growth retardation that did not affect birth and normal life. Moreover, when the *Hs3st1*$^{-/-}$ mice were submitted to pathologic stimuli, they exhibited a strong antithrombin-mediated proinflammatory phenotype[172]. Indeed, HS3ST1 controls the $HS^{AT}$ production in endothelial cells, in where the enzyme plays a role in the regulation of the inflammo-modulatory activities of antithrombin[172]. In human, none *HS3ST1* pathogenic variant has been reported, in agreement with a non-essential role for normal development and physiology. However, HS3ST1 is centrally implicated in appropriate response to stimuli or injury in tissues or organs that express it[172]. Interestingly, in the human brain, genetic variability in *HS3ST1* carrying loci has been correlated with neurocognitive endophenotypes in a population at risk to develop mild cognitive impairment (MCI) and Alzheimer's disease[173-175]. Although the role of 3-*O*-sulfation in physiology is yet unknown, studies on 3S-HS binders have allowed identification of neuropilin-1, a heparin binding protein implicated in angiogenesis and axon guidance[176]. Interestingly, HS3ST1 activity seems not to be compensated by other HS3STs[177] likely because a requirement of specific substrate precursor pools in cells expressing several HS3STs[177]. Because the *HS3ST1* tissue restricted expression and as non-pathogenic variants are known in human, and as *Hs3st1*$^{-/-}$ mice live healthy and fertile (their light intrauterine growth retardation lacked pathological outcome) and requires stimuli to observe traits, *HS3ST1* was allocated to the Group IV of HSPG biosynthetic machinery genes (Fig. 4 and Fig. 5).

*HS3ST2* (Chr.16; ID: 9956)

*HS3ST2* codes for HS glucosamine 3-*O*-sulfotransferase 2 (HS3ST2), an enzyme predominantly expressed in brain and in less extent in lung and some other tissues[169] (Fig. 4). This protein belongs to the HS3ST family of enzymes that act during the last step of HS biosynthesis by transferring a sulfate group from PAPS to the C3 of GluA2S/IdoA2S-GlcNS±6S motifs to generates rare GluA2S/IdoA2S-GlcNS**3S**±6S motifs, which are different from those generated by HS3ST1[171]. 3S-HS carrying the rare IdoA2S-GlcNS3S±6S motive are called $HS^{gD}$ since they serve as receptors for HSV1 entry into cells through interaction with the gD protein on the virus capsid[171,178]. In human, non *HS3ST2* pathogenic variants have been reported, and *Hs3st2*$^{-/-}$ mice are viable, healthy, and fertile (under submission), suggesting the non-requirement of this enzyme for normal development and physiology. However, the absence of deleterious phenotypes in human and in the knockout mouse could be due to transcriptional and functional redundancy with other HS3STs that also produce $HS^{gD}$, including HS3ST3A, HS3ST3B, HS3ST4, HS3ST5, and HS3ST6[171], or because the enzyme activity is only relevant when an adequate stimuli is applied, as observed for other HS3STs[177]. Redundancy is supported by the fact that $HS^{gD}$ is made by several HS3STs, although the few data available in HS3ST2 function suggest its involvement in discrete stimuli-related biological events. For instance, it has been reported that *Hs3st2* expression increases in the rat pineal gland during daylight and decreases at night, when it can be re-induced by light stimuli through adrenergic/cAMP signaling[179]. Consistent with a role in brain, in where the enzyme is predominantly expressed, we have found that HS3ST2 is implicated in tauopathy development[180]. Interestingly, *HS3ST2* expression is regulated by methylation on the 5'UTR regions of its promoter, and controversy exists between a possible role as oncogene or as tumor suppressor[181]. Since *HS3ST2* shows tissue restricted expression, and as *Hs3st2* knockout mice develop normally, are viable and fertile, *HS3ST2* was allocated to the Group IV of HSPG biosynthetic machinery genes (Fig. 4 and Fig. 5).

*HS3ST3A1* (Chr.17 ; ID: 9955)

*HS3ST3A1* codes for HS glucosamine 3-*O*-sulfotransferase 3A1, a Golgian enzyme that transfers sulfate groups from PAPS to IdoA±2S-GlcN±S±6S to generate IdoA±2S-GlcN±S3**S**±6S in HS[171,169,]. *HS3ST3A1* shows partial tissue restricted expression with predominance in spleen (Fig. 4) and, although at lower extent, in reproductive tissues and in the glomerular basement membrane[177]. *Hs3st3a1*$^{-/-}$ mice have not yet been generated and non-pathogenic clinical variants have been reported in human, suggesting a non-essential role of this enzyme in normal development and homeostasis. However, *HS3ST3A1*, as other tissue restricted genes, might be implicated in the regulation of discrete physiological events that might occur after stimuli in the tissues in which the gene is expressed, for instance in placenta, in where *HS3ST3A1* expression increases during normotensive and pre-eclamptic pregnancies[182]. As other HS3STs, HS3ST3A1 has also been involved in HSV1 infection and genetic variability in the *HS3ST3A1* carrying locus has been associated to resistance to infection by *Plasmodium falciparum*[183,184], in agreement with the implication of 3S-HS during cells interactions with pathogens[171]. Similarly, as other HS3STs, *HS3ST3A1* expression is regulated by DNA methylation, and its association to malignant phenotypes has been proposed[181]. Since *HS3ST3A1* shows tissue restrictedly expression and no clinical variants are known in human, *HS3ST3A1* was allocated to the Group IV of HSPG biosynthetic machinery genes (Fig. 4 and Fig. 5).

*HS3ST3B* (Chr.17; ID: 9953)

HS glucosamine 3-*O*-sulfotransferase 3B1 (HS3ST3B1) is a Golgian enzyme that transfers 3-*O*-sulfates IdoA2S-GlcN±S±6S moieties in HS, generating IdoA2S-GlcN±S**3S**±6S sequences with non-anticoagulant activity, but that favor HSV1 infection[171]. *HS3ST3B* is expressed in several tissues with prevalence in liver, spleen, and lymph node, but lack in skeletal muscle, in where the highly restricted HS3ST5 is expressed (Fig. 4)[171]. *Hs3st3b1* knockout mice have not yet been generated, but a non-deleterious phenotype might be supposed as non-pathogenic variants have been reported in human. However, similarly to *HS3ST3A1, HS3ST3B1 expression* confers resistance to infection by *Plasmodium falciparum*[183-185] and HSV1 infection[171]. Moreover, a role of HS3ST3B1 during specific adaptative response to stimuli is suggested by its expression in peripheral T lymphocytes and Jurkat T cells. In these cells, cyclophilin B interacts with $HS^{gD}$ on responsive peripheral blood T lymphocytes, triggering migration and integrin-mediated adhesion[186]. Finally, as for HS3ST3A1, the enzyme affects tumor growth possible though an DNA methylation-related mechanism[181]. Because its restricted tissue expression, absence of clinical variants in human, and requirement for responsiveness to stimuli, *HS3ST3B1* was allocated to the Group IV of HSPG biosynthetic machinery genes (Fig. 4 and Fig. 5).

*HS3ST4* (Chr.16; ID: 9951)

Heparan sulfate-glucosamine 3-*O*-sulfotransferase 4 (HS3ST4) is a Golgian enzyme that transfers sulfate groups to C3 of IdoA2S-GlcNS±6S moieties in HS, generating IdoA2S-GlcNS**3S**±6S sequences with non-anticoagulant activity but which, thought production of $HS^{gD}$ sequences, favors HSV1 infection[171]. HS3ST4 is one of the HS biosynthetic enzymes with the higher tissue restricted expression[178], predominantly in brain, in spleen, and in some reproductive organs, as testis and prostate (Fig. 4). Although *Hs3st4* knockout mice have not yet been reported, its non-deleterious phenotype is suggested by the fact that in human non-pathogenic variants are known. However, GWAS suggest that genetic variability in *HS3ST4* represent susceptibility risk for age-related macular degeneration[187]. Similarly, in nondemented older people have shown *HS3ST4* SNP associations with verbal declarative memory, suggesting a role in synaptic function[188,189]. Accordingly with involvement in brain, variability on non-coding regions near the *HS3ST4* gene showed low but significant association with neuropathologic traits associated to tau protein burden in Alzheimer's disease[188]. Interestingly, we recently found that HS3ST4 is increased in Alzheimer's disease hippocampus[190]. Although the physiologic role HS3ST4 is still unknown, its tissue restricted expression suggest that it might play discrete functions, particularly in brain, in where it is predominantly expressed. As for other HS3STs, *HS3ST4* expression has also been observed in some cancer cells and its role as either tumor suppressor or oncogene is controverted[181]. Because of its strong tissue restricted expression and nonpathogenic variability in human, *HS3ST4* was allocated to the Group IV of HSPG biosynthetic machinery genes (Fig. 4 and Fig. 5).

*HS3ST5* (Chr.6; ID: 222537)

HS3ST5 is a HS biosynthetic enzyme of the HS3ST family that, as HS3ST4, shows high tissue restricted expression (Fig. 4), predominantly in the central nervous system[191], suggesting a role in neurologic functions. Interestingly, HS3ST5 is the only tissue restricted sulfotransferase expressed in skeletal muscle, although in very low extent. HS3ST5 produces $HS^{AT}$ and $HS^{gD}$ and has also been implicated in HSV1 infection[171]. *Hs3st5* knockout mice have not yet been reported and non-pathogenic *HS3ST5* mutations are known in human. However, GWAS have shown *HS3ST5* genetic polymorphism association with risk of microcephaly, cerebellar hypoplasia, and altered gray matter volume in patients with schizophrenia[192]. Because of its strong tissue restricted expression and non-pathogenic clinical variants, *HS3ST5* was allocated to the Group IV of HSPG biosynthetic machinery genes (Fig. 4 and Fig. 5).

*HS3ST6* (Chr.16; ID: 64711)

HS3ST6 is member of the type II membrane-bound protein HS3ST family of enzymes that show highly restricted expression. HS3ST6 predominantly expressed in skin, and in considerably lower extent in some reproduction organs, lung, urinary bladder, and colon (Fig. 4)[193]. Recently, whole exome sequencing studies have given evidence of *HS3ST6* SNP with traits characteristic of hereditary angioedema, which is a condition in which minor trauma or stress results in recurrent attacks of severe swelling of arms, legs, face, intestinal tract, and airways. Hereditary angioedema is characteristically due to mutations in *SERPING1*, a gene is involved in the regulation of the complement cascade, or in the *F12*, involved in the coagulation cascade. 3S-HS produced by HS3ST6 was recently proposed to be involved in the response to stimuli that triggers angioedema formation[194]. As *HS3ST6* is involved in response of tissue to stimuli, as trauma or stress, this gene was allocated to the Group IV of HSPG biosynthetic machinery genes (Fig. 4 and Fig. 5).

##### Group IV – Tissue restricted HS chain remodeling

*EXTL1* (Chr.1; ID: 2134)

*EXTL1* encodes the $\alpha$-1,4-*N*-acetylglucosaminyl transferase 1, also called exostosin like glycosyltransferase 1 (EXTL1), which catalyzes the addition of a GlcNAc to the non-reducing end of the growing HS chain, blocking HS chain elongation[132]. EXTL1 is the only EXTs/EXTL-type glycosyltransferase that shows tissue restricted expression, particularly in brain, spleen, muscle, and endometrium (Fig. 4). This might guaranty a stimuli-related discreet function in the tissue in which the gene is expressed. For instance, in spleen, the enzyme regulates the levels of HS, which likely function as a switch that controls the early maturation of B-cells, as shown in cultured B-cells of the transgenic mice overexpressing *Extl1*[195]; in that cells, the stimuli responsive effect was attributed to reduced levels on membrane associated HS in the overexpressing *Extl1* B-cell linage (heparin addition to cells had no effect). Moreover, genetic variability in this gene has been associated with muscle lipid composition suggesting a role in the regulation of lipid metabolism[196]. Besides, EXTL1 was also involved in the control of the growth of some cancer cells[197]. Although the knockout mouse has not been generated, no clinical variant has been reported for this gene, which is more likely involved in the adaptative response to stimuli, *EXTL1* was allocated to the Group IV of HSPG biosynthetic machinery genes (Fig. 4 and Fig. 5).
