## Supplementary material for "Variability in proteoglycan biosynthetic genes reveals new facets of heparan sulfates diversity. A systematic review and analysis": HSPG biosynthetic genes

### SUPPLEMENTARY TABLE

### THE HSPG BIOSYNTHETIC MACHINERY GENES

| Gene | Full name | NCBI gene ID | Chromosome | Location | Sequence : location | Also know as |
|---|---|---|---|---|---|---|
| ***AGRN*** | *agrin* | 375790 | *1* | 1p36.33 | NC_000001.11 (1020102..1056119) | CMS8; AGRIN; CMSPPD |
| ***B3GALT6*** | *beta-1,3-galactosyltransferase 6* | 126792 | *1* | 1p36.33 | NC_000001.11 (1232237..1235041) | ALGAZ; EDSP2; EDSSPD2; SEMDJL1; beta3GalT6 |
| ***B3GAT3*** | *beta-1,3-glucuronyltransferase 3* | 26229 | *11* | 11q12.3 | NC_000011.10 (62615296..62621986, complement) | JDSCD; GLCATI; glcUAT-I |
| ***B4GALT7*** | *beta-1,4-galactosyltransferase 7* | 11285 | *5* | 5q35.3 | NC_000005.10 (177600102..177610330) | XGPT; EDSP1; XGPT1; EDSSLA; XGALT1; EDSSPD1 |
| ***CANT1*** | *calcium activated nucleotidase 1* | 124583 | *17* | 17q25.3 | NC_000017.11 (78991716..79009793, complement) | DBQD; EDM7; DBQD1; SCAN1; SHAPY; SCAN-1 |
| ***COL18A1*** | *collagen type XVIII alpha 1 chain* | 80781 | *21* | 21q22.3 | NC_000021.9 (45405165..45513720) | KS; KNO; GLCC; KNO1 |
| ***EXT1*** | *exostosin glycosyltransferase 1* | 2131 | 8 | 8q24.11 | NC_000008.11: 117,762,756 - 118,143,560 | EXT, LGCR, LGS, TRPS2, TTV |
| ***EXT2*** | *exostosin glycosyltransferase 2* | 2132 | 11 | 11p11.2 | NC_000011.10: 44,079,906 - 44,267,624 | SOTV, SSMS |
| ***EXTL1*** | *exostosin like glycosyltransferase 1* | 2134 | 1 | 1p36.11 | NC_000001.11: 26,020,269 - 26,037,936 | EXTL |
| ***EXTL2*** | *exostosin like glycosyltransferase 2* | 2135 | 1 | 1p21.2 | NC_000001.11:100,870,068 - 100,897,715 | EXTR2 |
| ***EXTL3*** | *exostosin like glycosyltransferase 3* | 2137 | 8 | 8p21.1 | NC_000008.11:28,592,950 - 28,770,385 | RPR; BOTV; REGR; EXTR1; ISDNA; EXTL1L |
| ***GPC1*** | *glypican 1* | 2817 | *2* | 2q37.3 | NC_000002.12 (240435663..240468076) | glypican |
| ***GPC2*** | *glypican 2* | 221914 | *7* | 7q22.1 | NC_000007.14 (100169606..100177381, complement) | |
| ***GPC3*** | *glypican 3* | 2719 | *X* | Xq26.2 | NC_000023.11 (133535745..133985616, complement) | SGB; DGSX; MXR7; SDYS; SGBS; OCI-5; SGBS1; GTR2-2 |
| ***GPC4*** | *glypican 4* | 2239 | *X* | Xq26.2 | NC_000023.11 (133300103..133415489, complement) | KPTS; K-glypican |
| ***GPC5*** | *glypican 5* | 2262 | *13* | 13q31.3 | NC_000013.11 (91398621..92867237) | |
| ***GPC6*** | *glypican 6* | 10082 | *13* | 13q31.3-q32.1 | NC_000013.11 (93226807..94408020) | OMIMD1 |
| ***GLCE*** | *glucuronic acid epimerase* | 26035 | 15 | 15q23 | NC_000015.10: 69,149,389 - 69,283,375 | HSEPI |
| ***HPSE*** | *heparanase* | 10855 | 4 | 4q21.23 | NC_000004.12: 83,288,192 - 83,339,422 | HPA; HPA1; HPR1; HSE1; HPSE1 |
| ***HPSE2*** | *heparanase 2* | 60495 | *10* | 10q24.2 | NC_000010.11 (98457077..99235875, complement) | UFS; HPA2; HPR2; UFS1 |
| ***HS2ST1*** | *heparan sulfate 2-O-sulfotransferase 1* | 9653 | 1 | 1p22.3 | NC_000001.11: 86,895,099 - 87,129,534 | dJ604K5.2 |
| ***HS3ST1*** | *heparan sulfate-glucosamine 3-sulfotransferase 1* | 9957 | 4 | 4p15.33 | NC_000004.12: 11,389,025 - 11,438,523 | 3OST; 3OST1 |
| ***HS3ST2*** | *heparan sulfate-glucosamine 3-sulfotransferase 2* | 9956 | 16 | 16p12.2 | NC_000016.10: 22,803,944 - 22,926,556 | 30ST2; 3OST2 |

| | | | | | | |
|---|---|---|---|---|---|---|
| ***HS3ST3A1*** | *heparan sulfate-glucosamine 3-sulfotransferase 3A1* | 9955 | 17 | 17p12 | NC_000017.11: 13,483,238 - 13,612,768 | 3OST3A1; 3-OST-3A |
| ***HS3ST3B1*** | *heparan sulfate-glucosamine 3-sulfotransferase 3B1* | 9953 | 17 | 17p12 | NC_000017.11: 14,296,214 - 14,354,242 | 3OST3B1; 3-OST-3B; h3-OST-3B |
| ***HS3ST4*** | *heparan sulfate-glucosamine 3-sulfotransferase 4* | 9951 | 16 | 16p12.1 | NC_000016.10: 25,647,386 - 26,182,261 | 30ST4; 3OST4; 3-OST-4; h3-OST-4 |
| ***HS3ST5*** | *heparan sulfate-glucosamine 3-sulfotransferase 5* | 222537 | 6 | 6q21-q22.1 | NC_000006.12: 114,026,901 - 114,371,118 | 3OST5; 3-OST-5; HS3OST5; NBLA04021 |
| ***HS3ST6*** | *heparan sulfate-glucosamine 3-sulfotransferase 6* | 64711 | *16* | 16p13.3 | NC_000016.10 (1911475..1921428, complement) | 3OST6; HS3ST5; 3-OST-6 |
| ***HS6ST1*** | *heparan sulfate 6-O-sulfotransferase 1* | 9394 | 2 | 2q14.3 | NC_000002.12: 128,260,141 - 128,324,207 | HH15; HS6ST |
| ***HS6ST2*** | *heparan sulfate 6-O-sulfotransferase 2* | 90161 | X | Xq26.2 | NC_000023.11: 132,592,471 - 132,994,934 | MRXSPM |
| ***HS6ST3*** | *heparan sulfate 6-O-sulfotransferase 3* | 266722 | *13* | 13q32.1 | NC_000013.11 (96090107..96839562) | HS6ST-3 |
| ***HSPG2*** | *heparan sulfate proteoglycan 2* | 3339 | *1* | 1p36.12 | NC_000001.11 (21822244..21937310, complement) | PLC; SJA; SJS; HSPG; SJS1; PRCAN |
| ***NDST1*** | *N-deacetylase and N-sulfotransferase 1* | 3340 | 5 | 5q33.1 | NC_000005.10: 150,478,540 - 150,565,454 | HSST; NST1; MRT46 |
| ***NDST2*** | *N-deacetylase and N-sulfotransferase 2* | 8509 | 10 | 10q22.2 | NC_000010.11: 73,800,917 - 73,812,843 | NST2; HSST2; N-HSST 2 |
| ***NDST3*** | *N-deacetylase and N-sulfotransferase 3* | 9348 | 4 | 4q26 | NC_000004.12: 118,010,760 - 118,281,507 | HSST3 |
| ***NDST4*** | *N-deacetylase and N-sulfotransferase 4* | 64579 | 4 | 4q26 | NC_000004.12: 114,795,121 - 115,143,134 | N-HSST; NDST-4; NHSST4; N-HSST 4 |
| ***PXYLP1*** | *2-phosphoxylose phosphatase 1* | 92370 | *3* | 3q23 | NC_000003.12 (141231825..141294924) | XYLP; ACPL2; HEL124 |
| ***SDC1*** | *syndecan 1* | 6382 | *2* | 2p24.1 | NC_000002.12 (20200797..20225475, complement) | SDC; CD138; SYND1; syndecan |
| ***SDC2*** | *syndecan 2* | 6383 | *8* | 8q22.1 | NC_000008.11 (96493601..96611790) | HSPG; CD362; HSPG1; SYND2 |
| ***SDC3*** | *syndecan 3* | 9672 | *1* | 1p35.2 | NC_000001.11 (30869466..30909735, complement) | SDCN; SYND3 |
| ***SDC4*** | *syndecan 4* | 6385 | *20* | 20q13.12 | NC_000020.11 (45325288..45348424, complement) | SYND4 |
| ***SLC10A7*** | *solute carrier family 10 member 7* | 84068 | *4* | 4q31.22 | NC_000004.12 (146253981..146522351, complement) | P7; SSASKS; C4orf13 |
| ***SULF1*** | *sulfatase 1* | 23213 | 8 | 8q13.2-q13.3 | NC_000008.11: 69,447,195 - 69,680,341 | SULF-1 |
| ***SULF2*** | *sulfatase 2* | 55959 | 20 | 20q13.12 | NC_000020.11: 47,641,363 - 47,799,835 | HSULF-2 |
| ***FAM20B*** | *FAM20B glycosaminoglycan xylosylkinase* | 9917 | 1 | 1q25.2 | NC_000001.11 (179025804..179076567) | gxk1 |
| ***XYLT1*** | *xylosyltransferase 1* | 64131 | *16* | 16p12.3 | NC_000016.10 (17101769..17470960, complement) | XT1; XTI; XT-I; DBQD2; XYLTI; PXYLT1; xylT-I |
| ***XYLT2*** | *xylosyltransferase 2* | 64132 | *17* | 17q21.33 | NC_000017.11 (50346126..50361185) | SOS; XT2; XT-II; PXYLT2; xylT-II |
